# Wearable Data Reveals distinct Characteristics of Individuals with Persistent Symptoms after a SARS-CoV-2 Infection

**DOI:** 10.1101/2024.05.21.24307673

**Authors:** Katharina Ledebur, Marc Wiedermann, Christian Puta, Stefan Thurner, Peter Klimek, Dirk Brockmann

## Abstract

Evidence on long-term health effects of SARS-CoV-2 infection remains scarce. Using a wearable-derived behavioral and physiological dataset (n = 20,815), we identified 137 individuals characterized by self-reported persistent fatigue and shortness of breath following a SARS-CoV-2 infection. Compared with symptom-free COVID-19 positive and negative controls, these individuals had higher resting heart rates (mean difference 2.37/1.49 bpm) and lower daily step counts (mean 3,030/2,909 steps fewer), even at least three weeks *prior* to SARS-CoV-2 infection. In addition, persistent COVID-19 symptoms were associated with a significant reduction in mean quality of life (WHO-5, EQ-5D), even *before* infection. Analysis of individual wearable time series suggests that individuals with persistent symptoms may have been more exposed to pre-existing health conditions/lower fitness levels prior to SARS-CoV-2 infection. Our study demonstrates the potential of wearable devices to track physiological and physical activity dynamics under natural conditions in the context of infectious and chronic diseases.

## Introduction

SARS-CoV-2 infections exhibit a diverse range of long-term impacts on individual health. The WHO defines post-COVID-19 condition (PCC) as *the continuation or development of new symptoms 3 months after the initial SARS-CoV-2 infection, with these symptoms lasting for at least 2 months with no other explanation* [1]. PCC is a multi-systemic condition [2] that has led to increased demand for health care due to increased disease burden [3]. Studies on persistent symptoms post-SARS-CoV-2 infection vary widely in design, including differences in symptoms studied, follow-up duration, and study populations [4–7]. In addition, challenges in data availability - such as a lack of adequate control cohorts, insufficient pre-infection health data, a focus on severe cases, and a reliance on subjective self-reports or infrequent measurements - have limited our understanding of persistent symptoms post-SARS-CoV-2 and those affected by it [5, 8–12].

Data from wearable devices such as smartwatches or fitness trackers, offer a novel approach to understanding the long-term impact of SARS-CoV-2 by providing objective measurements of heart rate, physical activity, step count, and sleep duration [13–15]. Combining this data with individual symptom reports not only enables the detection of critical events (e.g infection onset) but also also provides a deeper understanding of how consequences of a SARS-CoV-2 infection affect individual physiology and behavior [16–18]. These datasets provide a unique opportunity to establish pre-infection health baselines through continuous, non-invasive, and cost-effective measurements.

During the COVID-19 pandemic, several initiatives were launched to collect such data [18–20]. One such initiative is the Corona Data Donation project [13, 21–23] by the Robert Koch Institute, Germany’s federal agency for public health research. Between April 2020 and December 2022, over 500,000 voluntary participants registered for the project, obtained and activated the Corona Data Donation Application (CDA), and over 120,000 submitted more than 600 days of their basic wearable data (such as resting heart rate (RHR) and step count) using a custom-made smartphone application. In addition to daily averages, wearable data is available with exceptional resolution, down to the order of seconds. This permits a deep analysis of wearable-derived time-series, e.g. short high-intensity activities, regularities, and potential disruptions under natural conditions. Moreover, the high temporal resolution of the data and its coverage of long periods of time permit a detailed analysis of wearable data signals in all phases of a SARS-CoV-2 infection, i.e. the acute phase (from the week of the positive SARS-CoV-2 test to four weeks after the test), the sub-acute phase (four weeks to 12 weeks after the test), and the post-acute phase (12 weeks after the test date) [24]. One can clearly distinguish between individuals exhibiting weak regulation responses following infection (Fig. 1a,b) from those with strong ones (Fig. 1c,d). Participants also completed monthly and weekly surveys about COVID-19 tests, symptoms as well as their subjective wellbeing and health-related Quality of Life (QoL).

**Fig 1.**
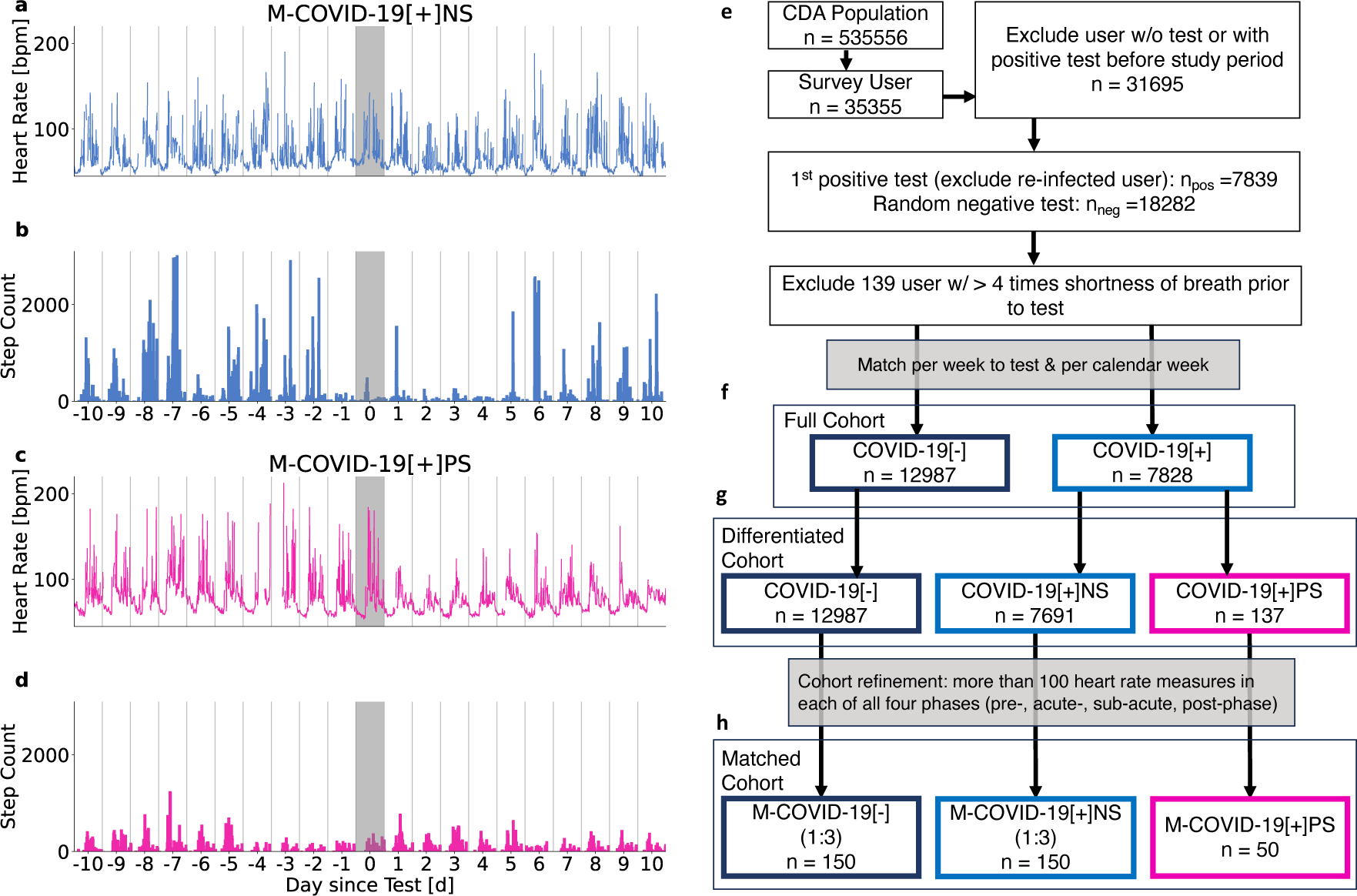
Time series of two representative individuals from the CDA population (a-d) and cohort diagram (e-h). Average daily heart rate [bpm] (a,c) and step counts (b,d) per 15 minutes relative to the day of the reported positive SARS-CoV-2 test (shaded grey area). Heart rate and step count for an individual without persistent symptoms 10 days before and after a reported positive test (a-b). Response in RHR and step count is more pronounced for an individual reporting persistent shortness of breath and fatigue (c-d). Both participants exhibited reduced step count a day prior to the test and in the following days, with the participant reporting persistent symptoms showing prolonged reduction. (e) Workflow of cohort creation. (f) Full cohort encompassing a positive cohort (*COV ID -* 19[+]) and negative control cohort (*COV ID -* 19[*-*]). (g) *Differentiated cohort* encompassing the persistent symptoms cohort (*COV ID -* 19[+]*PS*), the positive control cohort (*COV ID -* 19[+]*NS*), and the negative control cohort (*COV ID -* 19[*-*]). (h) *Matched cohort* consisting of the persistent symptoms match cohort (*M - COV ID -* 19[+]*PS*), the positive match cohort (*M - COV ID -* 19[+]*NS*) and the negative match cohort (*M - COV ID -* 19[*-*]).

We leveraged the unique combination of wearable data and survey data from the Corona Data Donation project to analyze the physical health of individuals reporting persistent symptoms following a SARS-CoV-2 infection. We focused on cohort-level averages of physiological metrics (specifically RHR) behavioral (step count) metrics. To make sense of the observed changes in wearable data, we then compared these objective measures with self-reported QoL (WHO-5 and EQ-5D) to show how signals in passively measured health data translate directly into subjectively experienced changes or limitations in daily life.

Our study aims to investigate resting heart rate, step count, subjective symptoms report and self reported quality of life to analyse possible interactions with pre-existing health conditions before infection, time-course of infection and post-infection phases. To this end, we first identified a cohort of individuals that reported a positive SARS-CoV-2 test (*COV ID -* 19[+]) along with their negative controls (*COV ID -* 19[*-*]). Within *COV ID -* 19[+] we compared those that reported long-lasting symptoms (*COV ID -* 19[+]*PS*) with those that did not (*COV ID -* 19[+]*NS*) to characterize the PCC phenotype.

## Results

### Increased frequency of reports on prolonged symptoms after infection

We evaluated the relative frequency of the reported symptoms of shortness of breath and fatigue in the *full cohort* (Fig.2) in pre- (< 0 weeks to the reported COVID-19 test), acute- (0-4 weeks), subacute (5-12 weeks), and post-phase (>12 weeks) [24]. During the acute phase of infection, fatigue was reported as a symptom by up to 69.76% (SE = 0.54%) of individuals in *COV ID -* 19[+] and by 19.53% (SE = 0.46%) of individuals in *COV ID -* 19[*-*]. Fatigue was reported significantly more often in *COV ID -* 19[+] than in *COV ID -* 19[*-*] for at least 6 weeks starting from the week of the reported SARS-CoV-2 test, Fig. 2a. Shortness of breath was reported by up to 13.55% (SE = 0.51%) and 2.46% (SE = 0.23%) by individuals in *COV ID -* 19[+] and *COV ID -* 19[*-*], respectively. It was increasingly reported for 15 weeks in *COV ID -* 19[+] (Fig. 2b). The combination of both symptoms was reported by up to 12.2% (SE = 0.09%) and 1.84% (SE = 0.05%) by individuals in *COV ID-*19[+] and *COV ID -* 19[*-*] and at increased frequency in *COV ID -* 19[+] for a minimum of 11 weeks, see Fig. 2c.

**Fig 2.**
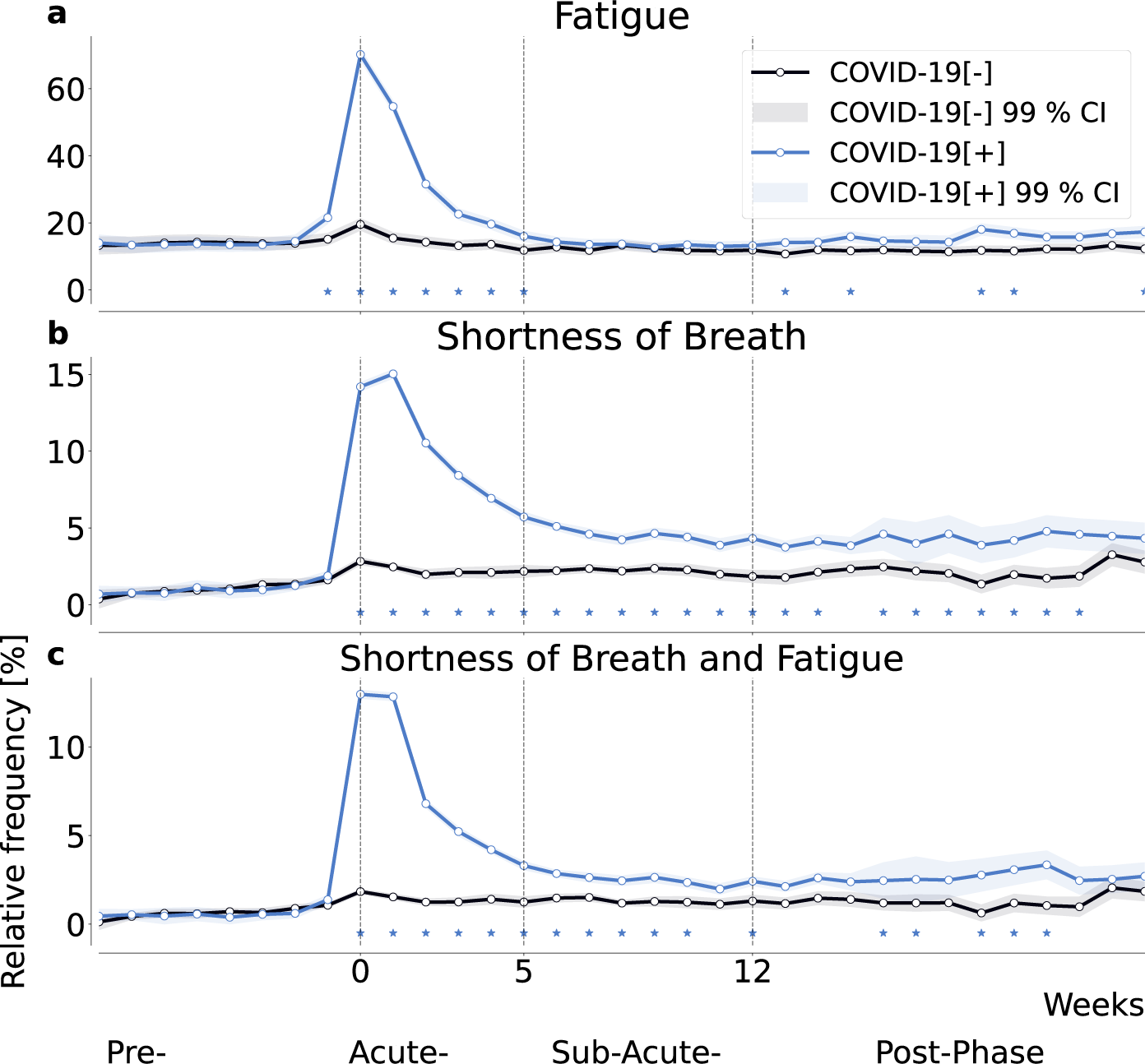
Relative frequency of self-reported fatigue (a), shortness of breath (b), and their combination (c), all relative to the week of the reported SARS-CoV-2 test for positive (P) individuals and a matched negative control (NC) cohort. Shading indicates the 99% confidence interval, i.e, 2.576 times the standard error of a binomial distribution. Asterisks indicate significant differences between the cohorts using a two-sided two proportion *z*-test with a significance level of 0.01.

Both, shortness of breath and fatigue, are potentially long lasting symptoms that could help identify individuals suffering from PCC [2, 25, 26]. Other assessed symptoms (cough, chills, loss of smell/taste, fever, diarrhea, limb ache and runny nose) did not significantly persist beyond the sub-acute phase when comparing *COV ID -* 19[+] with *COV ID -* 19[*-*] (Supplementary Figure 1).

Based on these findings, we defined a subset of *COV ID -* 19[+] as those individuals who reported shortness of breath and fatigue on at least 5 occasions from the week of the reported positive SARS-CoV-2 test (*COV ID -* 19[+]*PS*). Individuals in *COV ID -* 19[+]*PS* reported on average 6.2 (Std = 2.33, SE = 0.2) unique symptoms during the acute phase compared to 2.41 (Std = 2.39, SE = 0.01) symptoms reported by individuals with a positive SARS-CoV-2 test but no persistent symptoms (*COV ID-*19[+]*NS*) and 0.59 (Std = 1.14, SE = 0.00) symptoms reported by individuals in *COV ID-*19[*-*] (Supplementary Figure 2).

Supplementary Figure 3 illustrates relative frequencies of all symptoms in all phases, relative to the test date window for all cohorts, comprehensively.

### Persistent symptoms correspond to more pronounced and prolonged RHR-changes

We analyzed wearable-derived data around the SARS-CoV-2 test (Fig.3) for the three age- and gendermatched (1:3) cohorts: *M - COV ID -* 19[+]*PS*, *M - COV ID -* 19[+]*NS* and *M - COV ID -* 19[*-*] (Fig.1h).

**Fig 3.**
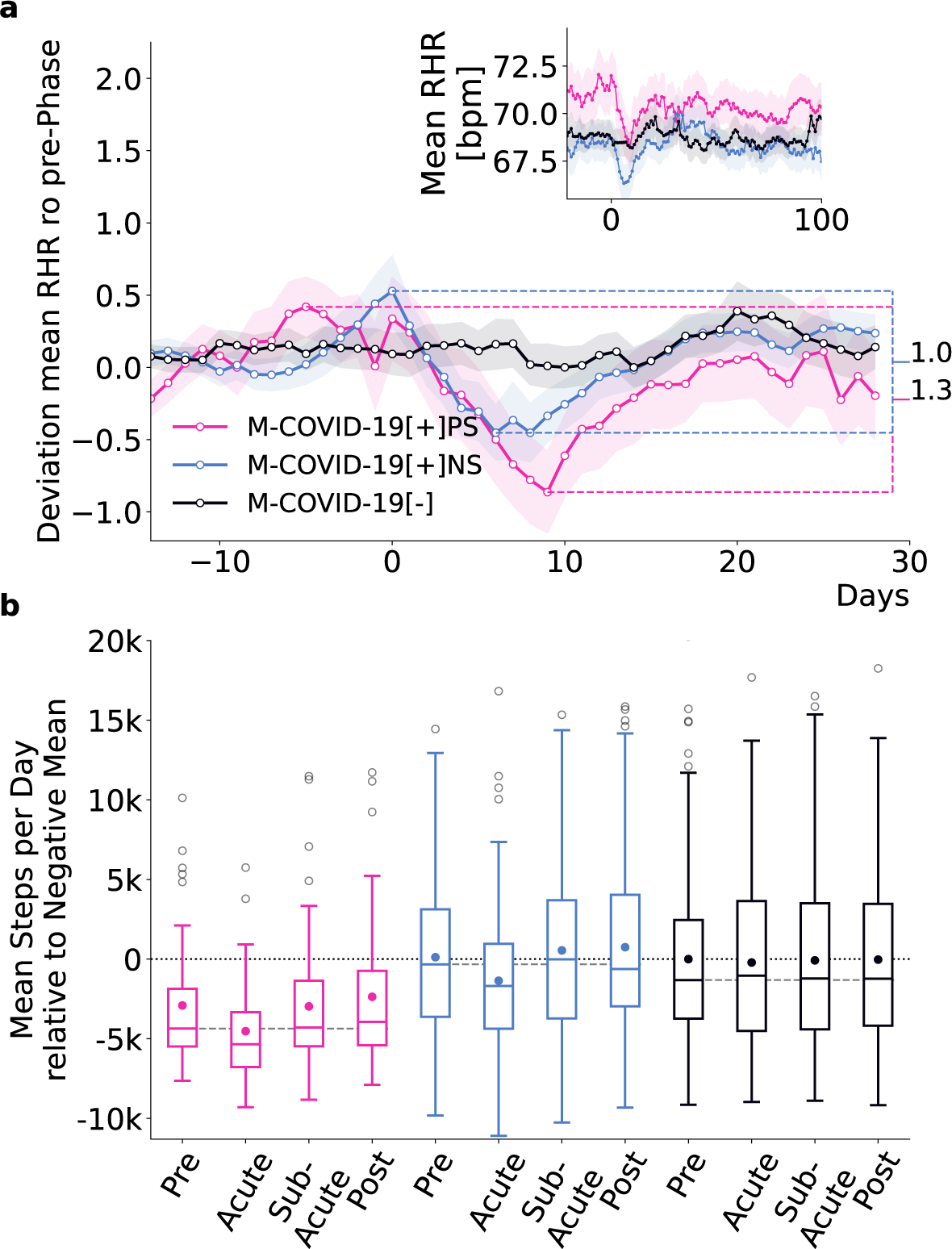
Wearable data analysis of the *match cohort*. (a) *Z*-transformed mean RHR (average of all 15-minute RHR measurements within the last seven days) relative to the seasonal mean RHR with respect to the mean and standard deviation up to seven days prior to the date of the reported test of all individuals in the *M - COV ID -* 19[+]*PS* (pink), *M - COV ID -* 19[+]*NS* (blue) and *M - COV ID -* 19[*-*] (black) cohorts. The difference between the maximum and minimum *z*-transformed RHR within 14 to and 20 days after the date of the reported SARS-CoV-2 test was more pronounced (1.3 vs 1.0) and more prolonged for *M - COV ID -* 19[+]*PS* than for *M - COV ID -* 19[+]*NS*. Shading indicates standard errors. The inset shows the average RHR relative to the SARS-CoV-2 test date. Already prior to the SARS-CoV-2 test, *M - COV ID -* 19[+]*PS*-individuals showed an increased RHR compared to *M - COV ID -* 19[+]*NS* and *M - COV ID -* 19[*-*]. (b) Average steps per day relative to the mean of *M - COV ID -* 19[*-*] during pre-phase (adjusted for seasonal variation) in all four phases for all individuals in all three cohorts. Boxes indicate quartiles, whiskers the range of the distribution of mean steps per day, scatter points outside the boxes mark the outliers (we do not show outliers > 20k steps per day), and scatter points within the box mark the mean. The dashed grey line indicates the median of the mean steps per day during the pre-phase. Median values for *M - COV ID -* 19[+]*PS* were consistently below the seasonal mean of the CDA population and below the median of the two control cohorts across all phases, indicating lower activity levels compared to the control cohorts. Mean number of steps per day for *M - COV ID -* 19[+]*PS* were below the mean number of steps per day of the two control cohorts in all four phases. Likewise we found a reduction in the variance of the *M - COV ID -* 19[+]*PS* compared to the two control cohorts.

To assess differences in RHR, we computed the average RHR for each individual over the last seven days, obtaining one data point per day relative to the test date. We adjusted for seasonal differences by subtracting the daily mean RHR of the CDA population (see Supplementary Figure 4a for seasonal differences in RHR).

To standardize the data and ensure comparability between individuals, we calculated per-user z-scores by subtracting each user’s average RHR and dividing by the standard deviation in the seven days prior to a reported SARS-CoV-2 test. This normalization centers all time series around zero, measured in units of standard deviation. We then averaged these Z-scores across all individuals within each cohort (see Methods for details).

We observed distinct RHR regulation patterns between the cohorts (Fig. 3 a). Individuals in *M - COV ID-* 19[+]*NS* and *M - COV ID-* 19[*-*] had comparable levels of mean RHR except during the acute phase.

The minimum Z-score during the acute phase for the *M - COV ID -* 19[+]*PS* cohort was -0.86, compared to -0.45 for the *M - COV ID -* 19[+]*NS* cohort, indicating that the average RHR in the *M - COV ID -* 19[+]*PS* cohort was 0.86 standard deviations below the pre-phase mean, compared to 0.45 standard deviations for the *M - COV ID -* 19[+]*NS* cohort. The range between the maximum and minimum deviation in the period 14 days before to 20 days after the test was more pronounced for the *M - COV ID -* 19[+]*PS* cohort, with a factor difference of 1.3 in the corresponding z-scores. In addition, *M - COV ID -* 19[+]*PS* individuals had a transient tachycardia followed by prolonged relative bradycardia, that did not return to baseline (*z*-score *2:* 0) until 18 days after infection. The relative bradycardia of *M - COV ID -* 19[+]*NS* individuals persisted until 15 days after infection.

Notably, *M - COV ID -* 19[+]*PS* individuals have on average an elevated RHR compared to the two control cohorts already *prior* to the reported SARS-CoV-2 test (inset in Fig.3a). Specifically, *M - COV ID -* 19[+]*PS* individuals exhibited an average increase of 2.37 bpm and 1.49 (between 21 to 7 days to the test) compared to *M - COV ID -* 19[+]*NS* and *M - COV ID -* 19[*-*] individuals, respectively.

### Physical activity profiles in acute and sub-acute infection regulation

To assess differences in physical activity patterns between individuals in the *M - COV ID -* 19[+]*PS*, *M - COV ID -* 19[+]*NS*, and *M - COV ID -* 19[*-*] cohorts, we analyzed the average step count per user per day in the pre-, acute-, sub-acute and post-phase of the *match cohort* (Fig.3b). Seasonal differences in activity (i.e., higher and lower step count during summer and winter, respectively, see (Supplementary Figure 4b)), were accounted for by subtracting the average of all participants in the CDA population from the average steps per day, see Methods Section for details. Results are presented relative to the mean steps per day during pre-phase of the *M - COV ID -* 19[*-*] cohort.

The respective median step count for *M - COV ID -* 19[+]*PS*-individuals was consistently below the median of both control cohorts in all four phases. Although the three cohorts are age- and sex-matched, the lower step counts in the *M - COV ID -* 19[+]*PS* cohort may reflect reduced activity levels. The mean step count per day was also below the mean step count of the two control cohorts in all four phases. During the pre-phase, the mean step count of the *M - COV ID -* 19[+]*PS* cohort was 3,030 steps less than that of the *M - COV ID -* 19[+]*NS* cohort and 2,909 steps less than the *M - COV ID -* 19[*-*] cohort. This indicates a generally lower level of activity compared to individuals from the *M-COV ID-*19[+]*NS* and *M-COV ID-*19[*-*] cohort. In addition, *M-COV ID-*19[+]*PS* individuals engaged at least three weeks prior to the infection in 1.12 and 0.99 days of high physical activity less than individuals from the *M - COV ID -* 19[+]*NS* and *M - COV ID -* 19[*-*] cohorts, respectively (Supplementary Figure 5).

### Persistent symptoms relate to lower well-being and quality of life

To explore the relationship between wearable data characteristics and perceived well-being and QoL, we analyzed responses to an adapted version of the WHO-5 wellbeing index [27, 28] and a modified EQ-5D health-related QoL survey (see SI for details) across the *differentiated cohort* (Fig. 1g), see Fig. 4. Due to the timing of the EQ-5D administration (post-2022), responses were not differentiated by pre- or post-SARS-CoV-2 test periods. By then, most participants had already reported a test (see Supplementary Figure 8).

**Fig 4.**
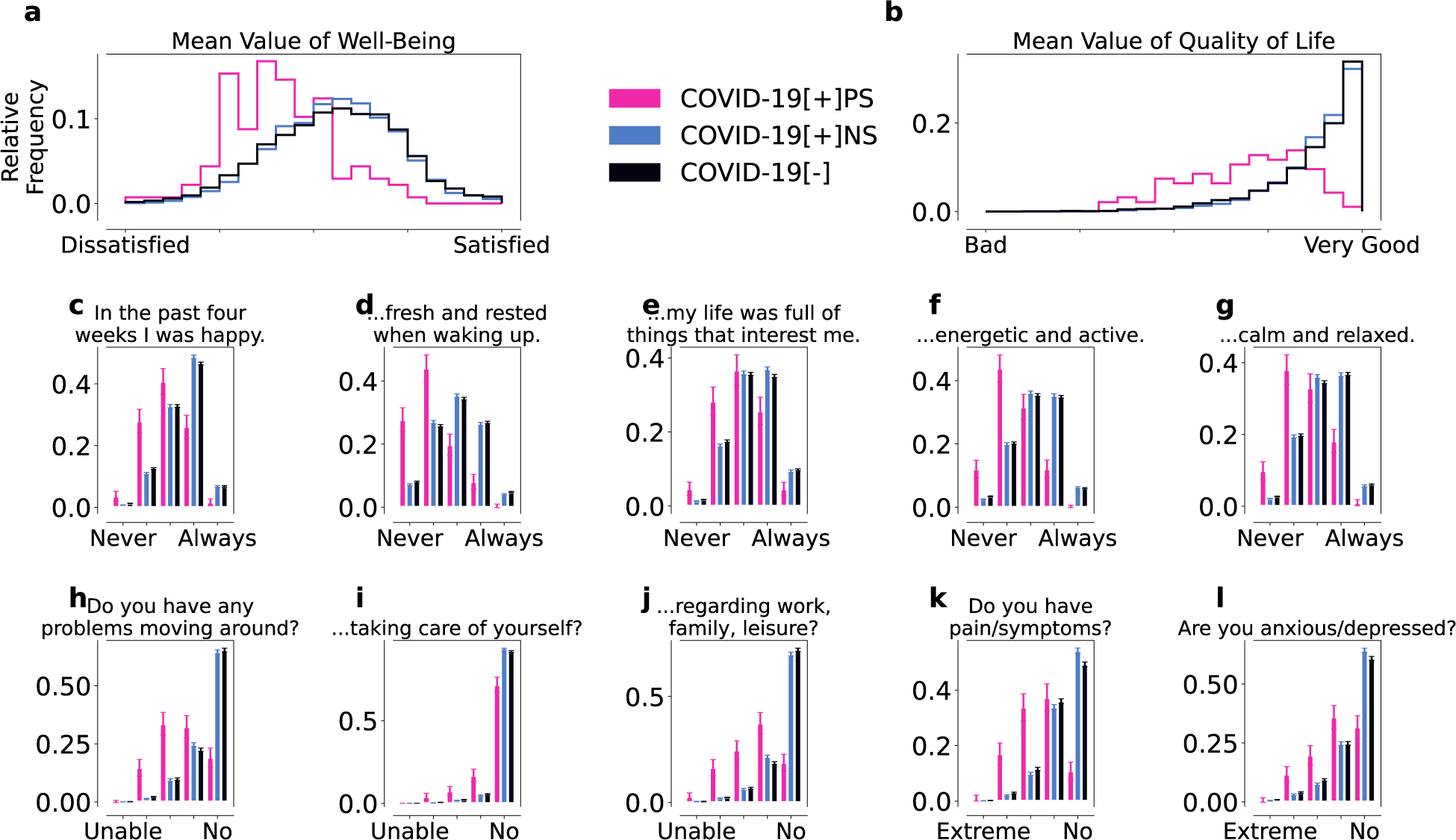
WHO-5 wellbeing (a,c-g) and modified EQ-5D/QoL (b,h-l) for the *COV ID -* 19[+]*PS* (pink), *COV ID -* 19[+]*NS* (blue), and *COV ID -* 19[*-*] (black) cohorts. The individual WHO-5 and modified EQ-5D scores were both averaged to obtain the overall wellbeing (a) and QoL (b) scores, respectively. Overall, *COV ID -* 19[+]*PS* individuals reported more issues with wellbeing (c-g) and QoL (h-l) than the control cohorts. Error bars indicate standard errors. The responses from all *COV ID -* 19[+]*PS* cohort were significantly different (*α <* 0.001) from the responses of the two control cohorts, as determined by the Mann-Whitney U test. All responses of the two control cohorts were significantly different as well except for the mean value of wellbeing (a), the mean value of QoL (b), and the questions “In the last four weeks I was calm and relaxed” (g) and “Do you have problems going around” (h).

Our version of the WHO-5 wellbeing index is constructed from five questions rated on a five-point scale (ranging from never (1) to always (5), with (3) is neutral; see SI for details). We examined the response for each individual question as well as the average response (Fig. 4 a, c-g). The overall distri-bution of wellbeing in *COV ID-*19[+]*PS* was skewed towards low values, (Fig. 4 a) with a mean value of 2.62 (Std = 0.55, SE = 0.05) for the *COV ID -* 19[+]*PS* cohort and a mean of 3.22 (Std = 0.63, SE = 0.01) and 3.2 (Std = 0.69, SE = 0.01) for the *COV ID -* 19[+]*NS* and *COV ID -* 19[*-*] cohort, respectively. We also observed lower wellbeing scores for all individual outcomes (Fig. 4 c-g). Users in *COV ID -* 19[+]*PS* reported particularly low scores for feeling energetic and rested upon waking up (Fig. 4d), possibly indicating activity and sleep issues. Statistically significant differences (*α* = 0.001) between the *COV ID-*19[+]*PS* and the control cohorts were found in regards to all WHO-5 questions, as determined by the Mann-Whitney U test. Likewise, we found statistically significant differences between the reports of the two control cohorts except for the questions “In the past four weeks I felt fresh and rested when waking up” (d), “In the past four weeks I was calm and relaxed” (g), and the mean of wellbeing. We observed statistically significant differences between all three cohorts, nevertheless the significance level (p-value) was lower when comparing the *COV ID -* 19[+]*PS* cohort to the control cohorts than for the intra-control cohort comparison.

In addition, we examined EQ-5D health-related QoL assessed by five questions answered on an ordinal scale ranging from the best (“I have no problems/No”; coded as 5) to the worst outcome (“I am not able to do that/extreme”; coded as 1) (Fig. 4b, h-l). The average response to the five questions, a measure of overall QoL, was systematically lower for individuals in *COV ID -* 19[+]*PS* compared to the control cohorts (Fig. 4b) with a mean of 2.79 (Std = 0.62, SE = 0.06). Again, *COV ID -* 19[+]*NS* and *COV ID -* 19[*-*] show similar values across modified EQ-5D questions and overall with a mean of 3.55 (Std = 0.46, SE = 0.01) and 3.52 (Std = 0.52, SE = 0.01) for *COV ID -* 19[+]*NS* and *COV ID -* 19[*-*], respectively. Likewise, *COV ID -* 19[+]*PS*-individuals reported lower QoL across all five questions (Fig. 4h-l) compared to the control cohorts, particularly concerning problems moving around and pain and physical symptoms (Fig. 4 b and k), again possibly indicating the presence of comorbidities or pre-existing conditions.

The answers of the *COV ID -* 19[+]*PS* cohort were significantly different from the control cohorts for all QoL questions (*α* = 0.001). Notably, also the answers of the *COV ID -* 19[+]*NS* compared to the *COV ID -* 19[*-*] cohort were significantly different except for the the mean of QoL (b) and the question “Do you have problems moving around?” (h), and “Do you have problems when it comes to general activities (work, studying, housework, family- or leisure activities)?” (j). However, the significance level was again lower when comparing the *COV ID -* 19[+]*PS* cohort to the two control cohorts than for the intra-control cohort comparison.

We also analyzed the responses to the wellbeing and QoL questions before (Supplementary Figure 6) and after (Supplementary Figure 7) the reported SARS-CoV-2 tests and found systematically lower values of wellbeing and QoL for the *COV ID -* 19[+]*PS* cohort before as well as after the reported test compared to the *COV ID -* 19[+]*NS* and *COV ID -* 19[*-*] cohorts. This suggests that individuals with lower levels in wellbeing and QoL may be at higher risk of developing long-term symptoms, possibly due to pre-existing general physical or mental health problems that may be related to another underlying condition.

In addition, we performed simple logistic regression analysis to identify characteristics that might serve as predictors of the development of persistent symptoms. Given the imbalance between the *M - COV ID-* 19[+]*PS* and *M - COV ID-* 19[+]*NS* cohorts, we focused on the area under the precision-recall curve (AUPRC). Using 10-fold cross-validation with stratified sampling, the model achieved an AUPRC of 0.75, indicating moderate predictive performance. Feature importance analysis, based on mean coefficients and odds ratios across cross-validation folds, identified number of unique symptoms reported during the acute-phase of infection, chronic bronchitis, unspecified sex, and allergies as the most influential features, with mean odds ratios (OR) of 4.40, 1.65, 1.53, and 1.35, respectively. Mean pre-infection RHR also emerged as a notable predictor, with a mean OR of 1.31. These findings suggest that integrating wearable-derived data such as RHR with traditional clinical variables can enhance the identification of individuals at higher risk of developing persistent symptoms. This highlights the potential value of wearable technology in supplementing conventional approaches for early detection and management of PCC.

## Discussion

Our study comprehensively characterizes how persistent self-reported symptoms in acute and postacute COVID-19 infection relate to changes in resting heart rate and physical activity measured by consumer grade wearable sensors. This observational study provides a descriptive analysis of the variation in heart rate, physical activity, and self-reported health status among different cohorts, and highlights patterns that may be linked to persistent symptoms following SARS-CoV-2 infection. Based on RHR, step count, and survey-based QoL, we identified patterns associated with a phenotype of persistent COVID-19 symptoms compared to COVID-19 positive and negative controls. Individuals with this phenotype are characterized by persistent (sub-acute and post-phase) shortness of breath and fatigue, and tended to have elevated RHR levels and less physical activity *prior* to infection, as well as, on average, lower well-being and health-related QoL scores compared with positive and negative controls.

We found notable differences in the prevalence of fatigue, shortness of breath, and their cooccurrence in SARS-CoV-2 positive and negative individuals. Both symptoms have been reported as typical, persistent symptoms of COVID-19 which are common in individuals with PCC[2, 5, 6, 29]. Indeed, according to a WHO case definition, fatigue and dyspnoea (shortness of breath) are the main symptoms of PCC [30]. Our analysis confirmed that these symptoms were significantly more frequent in the positive cohort than in the controls, with a marked persistence beyond the acute phase. Other symptoms did not show the same prolonged pattern, highlighting the specificity of these particular symptoms in relation to PCC.

A comparison of the wearable data (RHR and step count) revealed that during the acute phase (0-4 weeks after positive SARS-CoV-2 test) of COVID-19, individuals who reported persistent symptoms exhibited (1) higher RHR before infection, (2) on average more pronounced bradycardia up to 18 days after the positive SARS-CoV-2 test, and (3) lower physical activity before during and after infection compared to positive controls. Similar differences have been observed in previous studies when comparing SARS-CoV-2 positive and negative individuals [13, 14]. Individuals that later reported persistent symptoms already exhibited an elevated average RHR (mean increase of 2.37 bpm/1.49 bpm) compared to positive/negative control cohorts prior to their SARS-CoV-2 testing. In addition, these individuals were less likely to engage in high-intensity activities (i.e. regular physical activity and/or training) and were generally less active than the two control cohorts. Specifically, their daily step count was on average 3,030/2,909 steps less and they engaged in high activity 1.12/0.99 days less during the pre-phase than the positive/negative controls. Given the known inverse relationship between RHR and physical activity [31, 32], these patterns suggest that lower physical fitness levels, possibly due to pre-existing conditions, could increase susceptibility to developing long-term symptoms.

When comparing the *COV ID -* 19[+] and *COV ID -* 19[*-*] cohorts and the *COV ID -* 19[+]*NS* and *COV ID -* 19[*-*] cohorts, we found significantly higher reported vaccination rates in the positive cohorts, but no significant difference in the proportion reporting no vaccination. This suggests that the observed difference in vaccination rates may be due to reporting bias rather than actual differences in health status.

The positive cohorts (*COV ID -* 19[+] and *COV ID -* 19[+]*NS*) were also generally younger than the negative cohort (*COV ID -* 19[*-*]). This age difference may lead to an underestimation of our results. There were also fewer individuals in the negative cohort who reported their gender, suggesting a potential reporting bias.

When comparing the *COV ID -* 19[+]*PS* (persistent symptoms) and *COV ID -* 19[+]*NS* (no persistent symptoms) cohorts, we observed a significantly higher proportion of women in the persistent symptom group, suggesting an association between female gender and the development of persistent symptoms, consistent with the existing literature [4, 33].

We observed a significantly higher BMI in the *COV ID-*19[+]*PS* cohort compared to the *COV ID-* 19[+]*NS* cohort. While this suggests that individuals with persistent symptoms (*COV ID -* 19[+]*PS*) may have a higher BMI, it is important to note that the number of participants who provided both weight and height information was too small to reliably include BMI in the logistic regression analysis.

The *COV ID -* 19[*-*] cohort reported fewer cases of “no disease in the past 12 months,” indicating more comorbidities compared to the positive cohorts. They had higher rates of conditions such as hypertension, elevated cholesterol, chronic bronchitis, and mental health diagnoses. This suggests that the negative cohort may be less healthy overall, which may cause our results to represent a lower bound of the true effect, as the negative cohort may be less healthy overall.

When comparing the *COV ID-* 19[+]*PS* cohort to the *COV ID-* 19[+]*NS* cohort, the *COV ID-*19[+]*PS* group reported more allergies, asthma, chronic bronchitis, mental illness, and “other conditions”. This suggests a higher burden of comorbidities among those with persistent symptoms. Similarly, in the matched cohorts (*M - COV ID -* 19[+]*PS* vs. *M - COV ID -* 19[+]*NS*), the *M - COV ID -* 19[+]*PS* cohort had higher rates of allergies, chronic bronchitis, and mental illness, supporting the notion that pre-existing health conditions may play a role in the development of persistent symptoms. This is in line with findings on the association of these conditions and the risk of developing PCC [34, 35].

It is important to note that our findings should not be interpreted in a way that increased RHR, lower step count, and pre-existing health conditions solely determine the manifestation of persistent symptoms. Instead they emphasize that individuals displaying these characteristics should be given extra attention and protection measures.

Our analysis of self-reported well-being and QoL showed significantly lower levels for individuals with persistent symptoms compared to the control cohorts, both before and after the reported SARS-CoV-2 test. This suggests the possibility of underlying physical or mental health problems, such as chronic conditions or allergies (see Supplemetary Table 1), that may predispose individuals to persistent symptoms. Specifically we find a mean difference of 0.6/0.58 in mean well-being score and a mean difference of 0.76/0.73 between *COV ID -* 19[+]*PS* and *COV ID -* 19[+]*NS*/*COV ID -* 19[*-*] individuals. This is in line with recent findings, where individuals reporting persistent symptoms also reported poorer mental health in comparison to a control group [36], as well as a study reporting disturbed sleep as an important risk factor for PCC [37].

In addition to our descriptive study, we analyzed the importance of variables associated with the risk of developing persistent symptoms in a predictive model. We found that the number of symptoms during the acute infection phase, chronic bronchitis, unspecified sex, allergies, and mean pre-infection RHR were among the most influential factors. Wearable-derived data may thus complement previously identified risk factors for the development of persistent symptoms [38, 39].

While “unspecified gender” emerged as a risk factor, this likely reflects differences in reporting behavior rather than biological risk. Individuals who do not report gender may differ in unmeasured characteristics, such as privacy concerns or health-seeking behaviors, that affect symptom reporting or health outcomes. Thus, this finding should be interpreted with caution, as it may reflect data completeness issues or participant behavior rather than a direct risk for developing persistent symptoms.

The use of wearable data allows detailed examination of activity and heart rate patterns, providing insight not only into how SARS-CoV-2 infection may affect behavior and physiology in the long term, but also into the health status of individuals prior to contracting the disease. The data provides distinct advantages for evaluating the general health state of individuals holistically, continuous, under natural conditions, and outside of healthcare facilities [40, 41]. Wearables thus have the potential to improve patient-centered care by empowering individuals to monitor their health and manage their symptoms more effectively [42, 43]. In addition, data donation projects allow us to investigate individuals across a broad spectrum of SARS-CoV-2 infection severity, since participants enter the study prior to an infection. This approach enables us to establish a healthy state baseline, based on measurements prior to an infection, for individuals without biasing the study towards specific infection severities. In other words, participants are not selectively enrolled into the study based on their expected severity of subsequent SARS-CoV-2 infection symptoms or outcomes. This is of particular significance in the context of PCC research, since this condition is not exclusively linked to severe COVID-19 cases [44].

Several factors should be considered when interpreting our results. The Corona Data Donation Project had an overrepresentation of males and an underrepresentation of adolescents and the elderly (65+), who are at higher risk for long-term consequences of SARS-CoV-2 [13, 45]. The study population is likely to be more health-conscious than the general population because of the association of wearable device use with health-related habits [46]. We did not explicitly account for vaccination status, as most participants are partially vaccinated or did not report their status (see Table 1). This, along with other cohort-specific characteristics, highlights the need to interpret our findings in the context of our study population, which may not be fully generalizable to other groups. We also did not differentiate between variants of concern (VoC), which may trigger different immune responses and heart rate regulation. However, most participants reported their SARS-CoV-2 test date in 2022, when the Omicron variant (B.1.1.529) was dominant in Germany.

**Table 1.** Characterization of the study cohorts as defined in Fig. 1. For additional information on the reported health states per cohort, see Tab. S1.

|  | COVID-19<br>[+]<sup>1</sup> | COVID-19<br>[-] | COVID-19<br>[+]NS<sup>2</sup> | COVID-19<br>[+]PS<sup>3</sup> | M-COVID-19<br>[-] | M-COVID-19<br>[+]NS<sup>4</sup> | M-COVID-19<br>[+]PS<sup>5</sup> |
| --- | --- | --- | --- | --- | --- | --- | --- |
| N | 7,691 | 12,987 | 7,669 | 137 | 150 | 150 | 50 |
| Vaccinated [%] | 90.0*** | 88.3 | 90.0*** | 88.3 | 91.3 | 90.0 | 88.0 |
| Not vaccinated [%] | 0.4 | 0.5 | 0.4 | 0.7 | 0.0 | 0.0 | 2.0 |
| Mean Age<br>(Std) | 52.4***<br>(11.9) | 55.0<br>(12.0) | 52.4***<br>(11.9) | 52.7<br>(11.7) | 53.6<br>(10.5) | 53.6<br>(10.5) | 53.6<br>(10.5) |
| Female [%] | 36.1** | 34.1 | 35.9** | 47.4** | 50.0 | 50.0 | 50.0 |
| Male [%] | 54.4 | 54.7 | 54.7 | 40.1** | 40.0 | 40.0 | 40.0 |
| Unspecified [%] | 9.8*** | 11.9 | 9.8*** | 11.7 | 10.0 | 10.0 | 10.0 |
| Mean BMI<br>(Std) | 26.2<br>(4.7) | 26.5<br>(5.1) | 26.2<br>(4.7) | 27.7*<br>(5.7) | 26.9<br>(6.2) | 25.5<br>(4.0) | 27.4<br>(4.6) |

<sup>1</sup> COVID-19[+] vs. COVID-19[-]; <sup>2</sup> COVID-19[+]NS vs. COVID-19[-]; <sup>3</sup> COVID-19[+]PS vs. COVID-19[+]NS; <sup>4</sup> M-COVID-19[+]NS vs. M-COVID-19[-]; <sup>5</sup> M-COVID-19[+]PS vs. M-COVID-19[+]NS. p-values: \*p < 0.05, \*\*p < 0.01, \*\*\*p < 0.001.

**Table 2.**
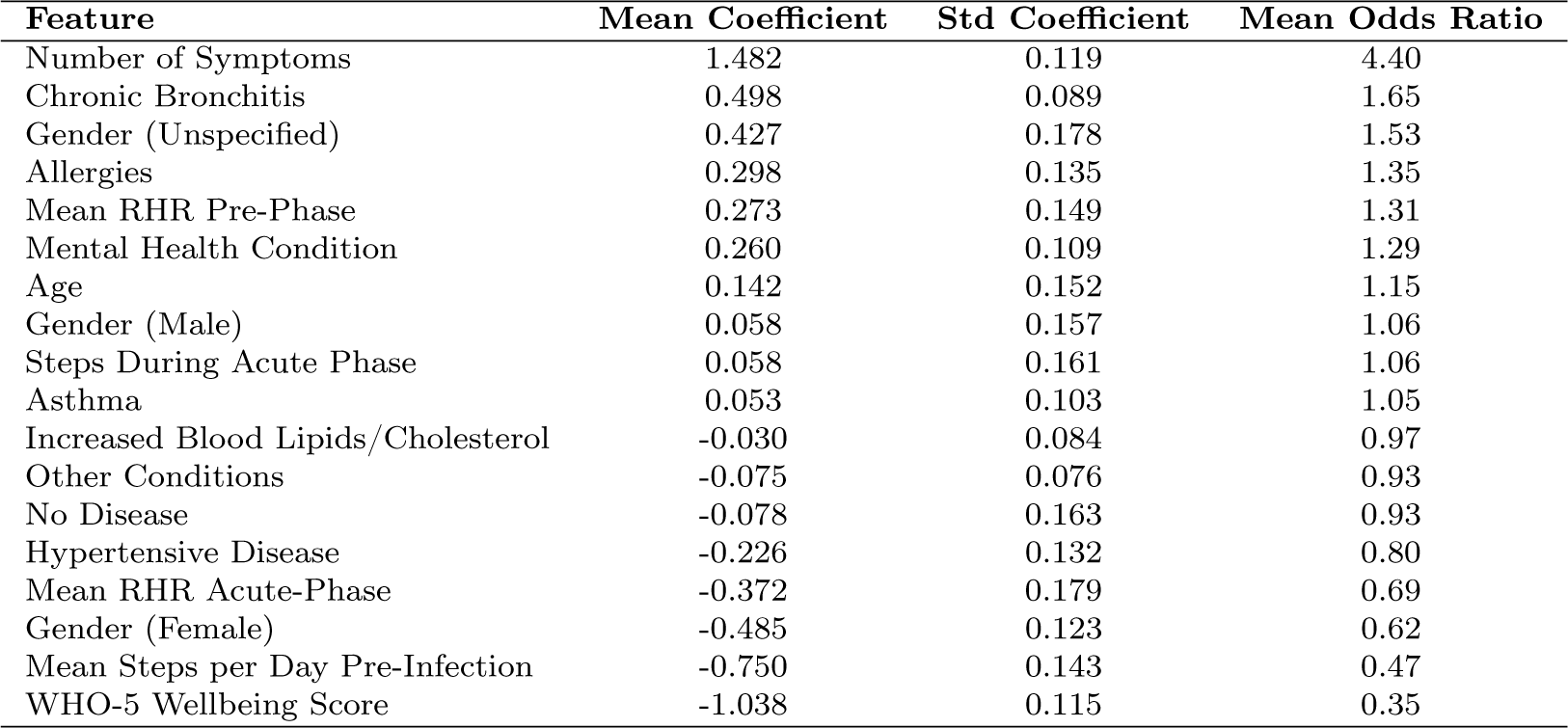
Predictive features for developing long COVID based on logistic regression analysis. The table shows the mean coefficients, standard deviations, and mean odds ratios from 10-fold cross-validation.

| Feature | Mean Coefficient | Std Coefficient | Mean Odds Ratio |
| --- | --- | --- | --- |
| Number of Symptoms | 1.482 | 0.119 | 4.40 |
| Chronic Bronchitis | 0.498 | 0.089 | 1.65 |
| Gender (Unspecified) | 0.427 | 0.178 | 1.53 |
| Allergies | 0.298 | 0.135 | 1.35 |
| Mean RHR Pre-Phase | 0.273 | 0.149 | 1.31 |
| Mental Health Condition | 0.260 | 0.109 | 1.29 |
| Age | 0.142 | 0.152 | 1.15 |
| Gender (Male) | 0.058 | 0.157 | 1.06 |
| Steps During Acute Phase | 0.058 | 0.161 | 1.06 |
| Asthma | 0.053 | 0.103 | 1.05 |
| Increased Blood Lipids/Cholesterol | -0.030 | 0.084 | 0.97 |
| Other Conditions | -0.075 | 0.076 | 0.93 |
| No Disease | -0.078 | 0.163 | 0.93 |
| Hypertensive Disease | -0.226 | 0.132 | 0.80 |
| Mean RHR Acute-Phase | -0.372 | 0.179 | 0.69 |
| Gender (Female) | -0.485 | 0.123 | 0.62 |
| Mean Steps per Day Pre-Infection | -0.750 | 0.143 | 0.47 |
| WHO-5 Wellbeing Score | -1.038 | 0.115 | 0.35 |

Keeping all these considerations in mind, it is reasonable to assume that our results under-represent the true disease burden of persistent symptoms and rather provide a lower bound for expected longterm imprints of an infection [36, 47–50]. We also did not control for wear time, but Supplementary Figure 9 shows similar distributions across cohorts.

The sample size of the study is limited by the requirement for sufficient wearable data (at least 100 heart rate measurements) in all four phases (pre, acute, sub-acute, and post) around a SARS-CoV-2 test. We also used strict criteria for defining persistent symptoms (at least five cases of shortness of breath and fatigue after the test), focusing on the most common symptoms observed in PCC [5–7, 29, 30]. While this focused approach inevitably introduces a degree of selection bias by targeting a specific subset of individuals with persistent symptoms, it is critical for accurately identifying those who are clearly affected and understanding their unique health profiles. We recognize that this targeted selection may limit the generalizability of our findings to broader populations. However, it allows for a more precise examination of the physiological and behavioral characteristics unique to those with persistent symptoms, thereby increasing the relevance and specificity of our findings.

The two cohorts studied for symptom reporting are not matched by age and gender, only by calendar week and time relative to a SARS-CoV-2 test. The small differences in age (2.6 years) and gender (1.5%) distributions between these cohorts are unlikely to affect the results(Table 1). Previous studies have found gender differences in symptoms only after the reported SARS-CoV-2 test for symptoms not included in our study [5].

Further studies are needed on the use of wearables to monitor physiological parameters and activity levels over time in individuals with PCC. Particularly, focusing on other persistent symptoms apart from shortness of breath and fatigue can aid in recognizing different patterns of symptom progression and immune response triggers. Moreover, high-frequency heart rate and activity can potentially be relevant for studying postural orthostatic tachycardia syndrome (POTS), a potential consequence of an infection with SARS-CoV-2 and a likely criterion for identifying PCC [51, 52]. Further combination of survey and wearable data would provide a deeper understanding of the relationship between subjective symptom reports and objective wearable data, allowing physiological and behavioural measures to be properly translated into experienced alterations to patients’ everyday lives. Further integration of wearable technology with telemedicine platforms facilitating the monitoring of symptoms in real-time and enabling remote consultation with healthcare professionals for individuals suffering from PCC can benefit patients and ease the burden on healthcare providers.

The unique advantage of wearable technologies is that continuous real-time data can be collected cheaply and on a large scale to provide fine-grained information on individual health, which is an ideal complement to traditional point-wise measurements in clinical research.

## Methods

### Data Characteristics

From April 12, 2020, to December 31, 2022, a total of 535,556 individuals downloaded and used the Corona-Datenspende App (CDA) [23], actively contributing a minimum of one essential data point each. The wearable data, i.e. RHR and step count, was submitted by linking the the app with consumer-grade smartwatches and fitness trackers. Participants could additionally consent to engage in periodic surveys on matters related to COVID-19, e.g., test results, weekly symptoms and monthly questions on QoL.

Weekly symptom reports were collected starting from October 21, 2021. By November 9, 2022, 35,355 individuals had participated in at least one survey. Participants could choose to report no symptoms or select one or more symptoms from a predefined list: Shortness of Breath, Fatigue, Headache, Sore Throat, Loss of Smell and Taste, Diarrhea, Runny Nose, Cough, Chills, Limb Ache, and Fever.

Comparisons were made between individuals who reported a positive SARS-CoV-2 test (*COV ID-* 19[+] = 7,691 individuals) and a negative control cohort (*COV ID -* 19[*-*] = 12,987 individuals) (see Methods and Fig.1e). Participants in the *COV ID -* 19[+] cohort reported symptoms an average of 18.4 times (standard deviation (Std) = 11.2). Participants in the NC cohort submitted reports 11.1 times on average (Std = 11.4). The *COV ID -* 19[+] cohort was further split in sub-cohorts, one with individuals that reported persistent symptoms (*COV ID -* 19[+]*PS* = 137 individuals) and the rest (*COV ID -* 19[+]*NS* = 7669 individuals), which together with the *COV ID -* 19[*-*] cohort yielding in three cohorts, see Methods and Fig.1e) for details on cohort creation.

For these cohorts we analysed survey responses on wellbeing and QoL. The questions on WHO-5 well-being were distributed since October 2021 and rolled out together with the symptom surveys. 35,492 individuals submitted corresponding answers. Particularly, all 137 individuals reporting persistent symptoms, 7,609 individuals from the *COV ID -* 19[+]*NS* cohort and 12,866 users from the *COV ID -* 19[*-*] cohort answered the survey on well-being at least once. The WHO-5 wellbeing index [27, 28] is answered on a six-point scale ranging from never(0) to always (5). For technical reasons the CDA uses a slightly altered version that utilizes a five-point scale ranging from 1 to 5 with 3 being neutral. The five questions ask: ‘In the past four weeks I was happy and in a good mood.’, ‘… I felt calm and relaxed.’, ‘… I felt energetic and active.’, ‘… I felt fresh and rested when waking up.’, and ‘… my everyday life was full of things that interest me.’

The modified EQ-5D survey on QoL was distributed since April 2022 to 12,179 participants. 92 individuals with persistent symptoms, 2,647 from the *COV ID-*19[+]*NS* cohort and 12,866 individuals from the *COV ID -* 19[*-*] cohort answered the five questions on QoL a least once. The survey asks to indicate what best describes the state of health today: ‘Do you have any problems moving around?’, ‘Do you have any problems taking care of yourself, washing or dressing yourself?’, ‘Do you have problems when it comes to general activities (work, studying, housework, family- or leisure activities)?’, ‘Do you have pain/physical symptoms?’, and ‘Are you anxious or depressed?’. The answers are on an ordinal scale ranging from the best possible outcome (“I have no problems/No”; coded as 5) to the worst possible outcome (“I am not able to do that/extreme”; coded as 1). For three age- and gender matched cohorts (1:3) denoted *persistent symptoms match* (*M - COV ID -* 19[+]*PS* = 50), *positive match* (*M -COV ID-*19[+]*NS* = 150) and *negative match* (*M -COV ID-*19[*-*] = 150), we analyzed high frequency longitudinal data on RHR and step count (see Methods and Fig. 1e). Data on RHR and step count were collected in intervals as low as 60 seconds. We show four exemplary timeseries spanning three weeks each in Fig.1a-d.

### Cohort creation

To study reported symptoms relative to a reported positive SARS-CoV-2 test (*COV ID -* 19[+]*PS*) and compared them with a negative control (*COV ID -* 19[*-*]) cohort we looked at 35,355 individuals who filled out the corresponding surveys. We excluded 3,660 individuals who reported a positive SARS-CoV-2 test in a previously distributed one-time questionnaire to avoid previous infections with SARS-CoV-2 in the population. We divided the population into two groups: those with at least one positive SARS-CoV-2 test and those with all negative tests. Both cohorts are limited to the period between October 21, 2021, and November 9, 2022. For individuals with positive tests, we used the date of their first positive test if multiple tests were positive. To avoid including individuals with re-infections, we excluded those reporting a test more than 28 days after the chosen test date, resulting in 7839 individuals. Among the 18,282 individuals reporting only negative tests, we randomly selected a test date. We also excluded users who reported shortness of breath more than four times before their test result, as a proxy for pre-existing respiratory conditions. To account for seasonal effects in symptom reporting, we adjusted the negative cohort to match the relative frequency of negative reports per calendar week and the temporal distance to the test week observed in the positive cohort. This yields a positive cohort (*COV ID -* 19[+]*NS*) of 7,691 individuals and a negative control (*COV ID -* 19[*-*]) of 12,987 individuals (full cohort, Fig. 1f).

For the analysis of WHO-5 and modified EQ-5D, we selected individuals from *COV ID-*19[+] who reported both shortness of breath and fatigue at least five times after their positive SARS-CoV-2 test. This subset, comprising 137 individuals, forms the persistent symptoms cohort (*COV ID -* 19[+]*PS*). We compared their survey responses with those of the remaining 7,669 individuals in the Positive Cohort (referred to as the positive control cohort (*COV ID -* 19[+]*NS*)) and the 12,987 individuals from the negative control (*COV ID -* 19[*-*]) cohort (differentiated cohort Fig. 1g).

To study differences and changes in physiological parameters (RHR) and behavior (step count) between the *COV ID -* 19[+]*NS*, *COV ID -* 19[*-*], and *COV ID -* 19[+]*PS* cohorts, we analyzed fine-grained wearable data collected by wearable devices. We standardized the data by aggregating high-frequency measurements into 15-minute intervals. Four phases relative to the reported SARS-CoV-2 test are defined: pre-phase (start of data collection until the week of the positive test), acute phase (zero to four weeks after the test), sub-acute phase (five to twelve weeks), and post-phase (twelve weeks or more after the positive test), aligning with recent definitions of COVID-19 disease stages [24]. After excluding individuals with less than 100 15-minute interval heart rate measurements across all phases, the persistent symptoms match (*M - COV ID -* 19[+]*PS*) cohort comprises 50 individuals. We created (1:3) age- and gender-matched positive match (*M - COV ID -* 19[+]*NS*) and negative match (*M -COV ID-*19[*-*]) cohorts, each with 150 individuals selected from the *COV ID-*19[+]*NS* and *COV ID -* 19[*-*] cohorts, respectively (match cohort, Fig. 1h). We chose a 1:3 matching, common in medical research, to increase statistical power and to improve the generalizability of our findings by ensuring that the control group more accurately represents the population from which the cases and controls were drawn.

For characteristics of the cohorts see Tabs. 1 and S1.

### Data processing and statistical analysis

#### Statistical analysis of group differences

To analyze group differences in our study cohorts, we used different statistical tests tailored to the nature of the data. Continuous variables, such as age and BMI, were analyzed using the Kruskal-Wallis test. Categorical variables, including sex, vaccination status, and comorbidities, were assessed using Fisher’s exact test.

We performed these comparisons in three main cohort analyses: In the *full cohort* analysis, we compared the positive and negative cohorts (*COV ID-*19[+] vs. *COV ID-*19[*-*]). In the *differentiated cohort* analysis, we compared the positive without persistent symptoms cohort and the negative cohort (*COV ID-*19[+]*NS* vs. *COV ID-*19[*-*]) as well as the positive with persistent symptoms cohort and the positive without persistent symptoms cohort (*COV ID -* 19[+]*PS* vs. *COV ID -* 19[+]*NS*). In the *matched cohort* analysis, we compared differences between the matched positive without persistent symptoms cohort and the matched negative cohort (*M -COV ID-*19[+]*NS* vs. *M -COV ID-*19[*-*]) and the matched positive with persistent symptoms cohort and the matched positive without persistent symptoms cohort (*M - COV ID -* 19[+]*PS* vs. *M - COV ID -* 19[+]*NS*).

For all statistical tests, significance levels were set at three thresholds: *α* = 0.05, *α* = 0.01, and *α* = 0.001.

#### Weekly reports on symptoms

For the *COV ID -* 19[+] and the *COV ID -* 19[*-*] cohorts, we compared the relative frequency of symptom reports relative to the calendar week in which a SARS-CoV-2 test was reported. We detected differences between the two groups at 99% confidence using a two-sided two proportion *z*-test and a significance level of 0.01. To investigate whether pairs of symptoms are significantly concordant, we compared the relative frequency of reports where individuals report both symptoms.

#### Resting heart rate and step count

To ensure data consistency, we only analyzed one source/device per participant (e.g., Apple Health, Fitbit, or Garmin). If a user has multiple devices, we prioritized the one with the most data points, potentially leading to missing information if different devices are used for different activities. Observations are based on measurement intervals ranging from less than one minute to several hours, resulting in overlapping intervals in a few instances and a slight overestimation of daily activity. This is rare compared to the total data volume. In our data pre-processing approach, intervals longer than one minute were split into one-minute segments and duplicate intervals are removed. These one-minute intervals were then resampled to 15-minute intervals by calculating the mean of the RHRs and the sum of the steps. In cases where a user contributes data from multiple sources, only the source with the highest number of entries is considered.

To compare RHRs between the *M - COV ID -* 19[+]*PS* cohort, the *M - COV ID -* 19[+]*NS* cohort and the *M - COV ID -* 19[*-*] cohort, for each user and each day we first computed (relative to the reported SARS-CoV-2 test) an average of all 15 minute intervals within the past seven days. We only computed the average if there were more than 20 entries within that seven-day window. We then computed the average RHR over all users in a cohort, again relative to the reported SARS-CoV-2 test. To evaluate tachycardia during the acute phase of the infection, we *z*-transformed the average of all 15 minute intervals within the last seven days per user relative to the mean RHR prior to the test (window of 62 days to 7 days before the date of the reported test). We additionally controlled for seasonal fluctuations by *z*-transforming the average RHR relative to the seasonal mean of the CDA population (Supplementary Figure 4a).

To examine differences in behavioral changes around a SARS-CoV-2 infection between cohorts, we investigated the average step count for the cohort members. As the daily step count shows seasonal variations [13], we always assessed this metric by subtracting the respective mean value per day and device within the entire set of participants of the whole Corona Data Donation project, see also Supplementary Figure 4b. We show the difference to the mean daily step count (adjusted for seasonal variation) of the *M - COV ID -* 19[*-*] cohort during pre-phase. A positive/negative value then indicates that an individual is more/less active than the *M - COV ID -* 19[*-*] cohort during pre-phase average. To assess variations or shifts in high-intensity physical activity, such as sports, we established a criterion where a day is considered to be high-intensity physical activity if the step count is more than one standard deviation above the seasonal average step count.

#### Monthly surveys on well-being and quality of life

For the analysis of the surveys on well-being/QoL we calculated the mean response to each question per user over all answers (not discriminating between before and after day of reported SARS-CoV-2 test) and showed the relative frequency of each possible answer value per cohort. Error bars indicate the standard error. The answers to each one of the five WHO-5 Well-Being questions are designed so that they can be averaged to obtain a mean value of well-being. Note, that the WHO-5 well-being is usually measured on a six-point scale. Due to technical reasons however, our survey provides an ordinal five-point scale. This also applies to the five modified EQ-5D QoL questions. The modification is attributed to different translations of questions and answers, along with one missing question. For both the WHO-5 and the modified EQ-5D, we show a histogram of the distribution of the respective mean scores for all three cohorts studied.

#### Logistic Regression

We performed logistic regression analysis to identify variables associated with the risk of developing persistent symptoms after SARS-CoV-2 infection. To ensure robustness, we used 10-fold stratified cross-validation to assess model performance and calculate the area under the precision-recall curve (AUPRC). Feature importance was assessed by calculating mean coefficients, standard deviations and odds ratios across cross-validation folds.

### Ethical consideration

Participation in the study was voluntary and self-recruited. All individuals participating in the Corona Data Donation Project provided informed consent electronically via the app. Consent was provided separately for submitting wearable data and participating in the in-app surveys. Participation is only possible for German residents age 16 and older and data is only stored pseudonymously, using a randomly generated unique user ID. Participant age is rounded to 5 years. The study is subject to strict compliance with the data protection provisions set out in the EU General Data Protection Regulation (GDPR) and the Federal Data Protection Act (BDSG). A comprehensive privacy impact assessment was conducted through an external law-firm specialized in e-Health and research projects. The study was reviewed and approved by the Data Privacy Officer at the Robert Koch Institute (internal operation number 2021-009) in agreement with the Federal Commissioner for Data Protection and Freedom of Information (BfDI), Germany’s highest independent supreme federal authority for data protection and freedom of information. Ethical approval for this study was obtained from the ethics board at the University of Erfurt (approval number 20220414).

## Data availability

The data analyzed in this study concerns the health condition of individual persons. As per the General Data Protection Regulation (GDPR) such data constitutes sensitive information and is protected as a “special category of personal data”. For this reason, the data must not be directly shared in a public repository, but interested parties can request access to the data following registration with the Data Privacy Officer at the Robert Koch-Institute. All initial data inquiries should be addressed to Dirk Brockmann or the general contact address of the Corona Data Donation project.

## Code availability

Python 3.11.6 was used to perform the simulations and data analysis. The simulation and analysis code for this study will be available under MIT license in the repository at https://github.com/566 katharinaledebur/characteristics_persistent_symptoms.git.

## Acknowlegdements

This study was financed with funds from the overall funding program of the City of Vienna MA7. Funding was also received from the Federal Ministry of Health of Germany (Grants “Corona-Datenspende”, CD21, DS22 and DS23).

## Author contributions

MW and KL conceptualized the study. MW supervised the project. DB devised and created the Corona Data Donation project. MW and KL devised the analytic methods. KL carried out the analysis and produced the plots and graphics. KL and MW directly accessed and verified the underlying data reported in the manuscript. KL and MW wrote the first draft of the manuscript. CP, DB, ST and PK made critical comments regarding the manuscript. CP contributed medical expertise regarding the medical interpretation of the findings and in developing medical hypotheses. KL, MW, CP, PK, DB and ST conducted reviewing and editing of the manuscript. All authors read and approved the final manuscript.

## SUPPLEMENTARY INFORMATION: WEARABLE DATA REVEALS DISTINCT CHARACTERISTICS OF INDIVIDUALS WITH PERSISTENT SYMPTOMS AFTER A SARS-COV-2 INFECTION

**Supplementary Table I.**
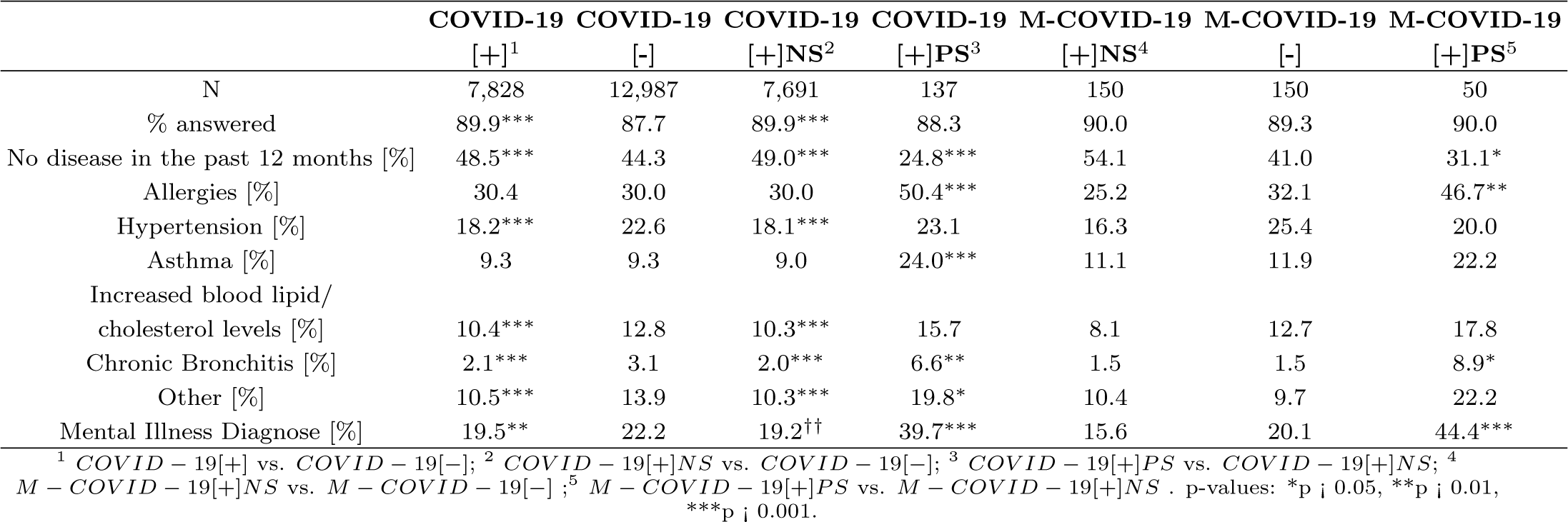
Information on other diseases and conditions of all studied cohorts. See Materials and Methods (Section IV) for further details on the specifics of the assignments.

**Supplementary Figure 1.**
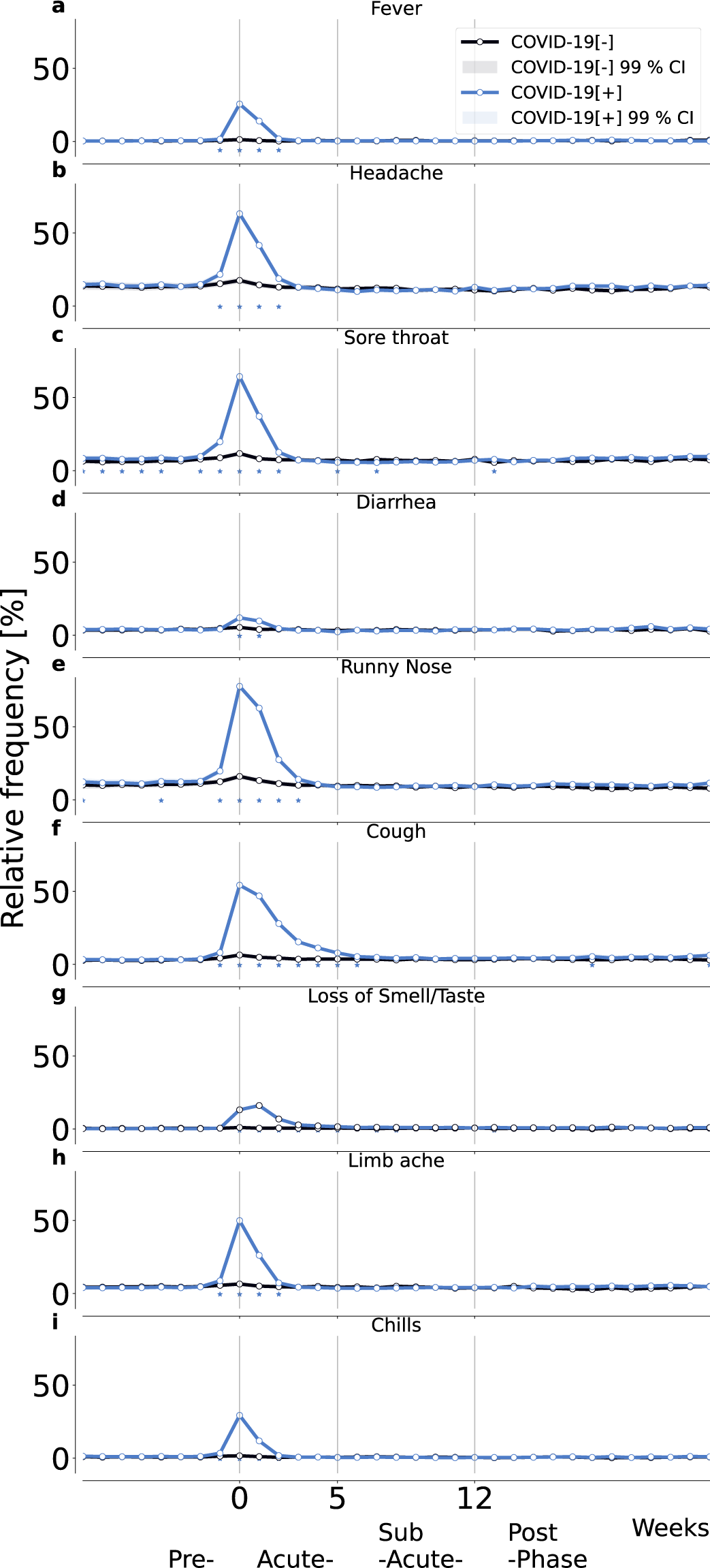
Relative frequency of symptoms relative to week of reported SARS-CoV-2 test for positive cohort in blue (*COV ID -* 19[+]) and negative control cohort (*COV ID -* 19[*-*]) in black. The symptoms are fever (a), headache (b), sore throat (c), diarrhea (d), runny nose (e), cough (f), loss of smell/taste (g), limb ache (h), and chills (i). Shading indicates the 99% confidence interval, i.e, 2.576 times the standard error of a binomial distribution. Asterisks indicate significant differences between the cohorts using a two-sided two proportion *z*-test with a significance level of 0.01.

**Supplementary Figure 2.**
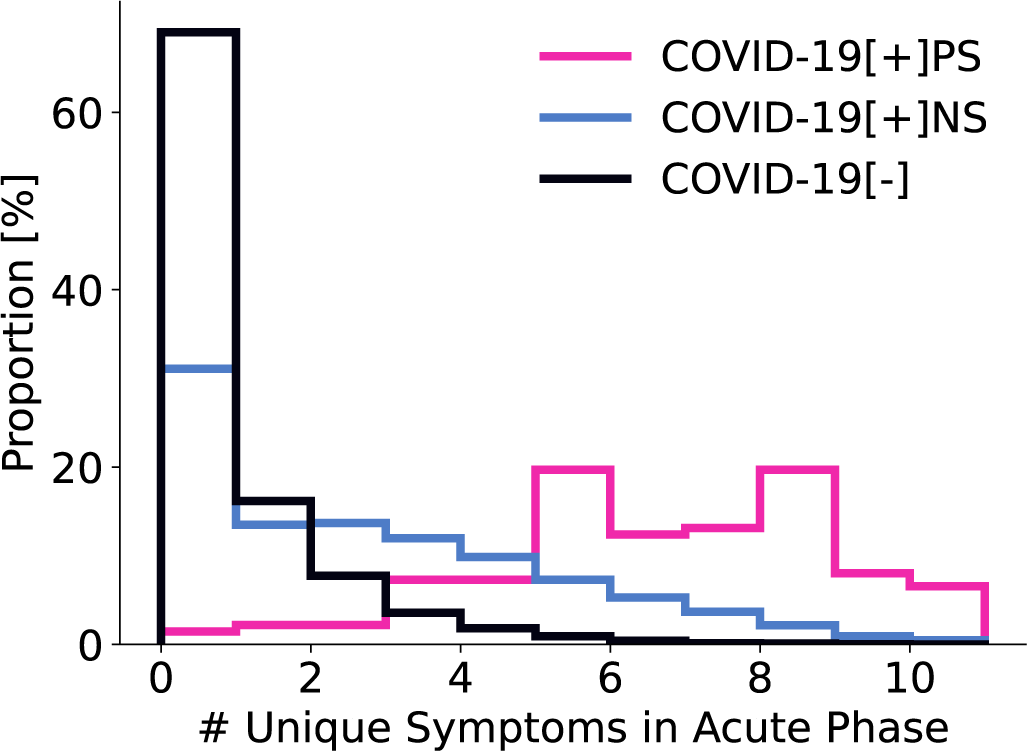
Distribution of number of unique reported symptoms during acute phase of the infection (0-4 weeks to the reported SARS-CoV-2 test). We show the proportion of individuals per number of unique symptoms during the acute-phase of the infection for the persistent symptoms in pink (*COV ID -* 19[+]*PS*), positive control in blue (*COV ID -* 19[+]*NS*) and negative control cohort in black (*COV ID -* 19[*-*]).

**Supplementary Figure 3.**
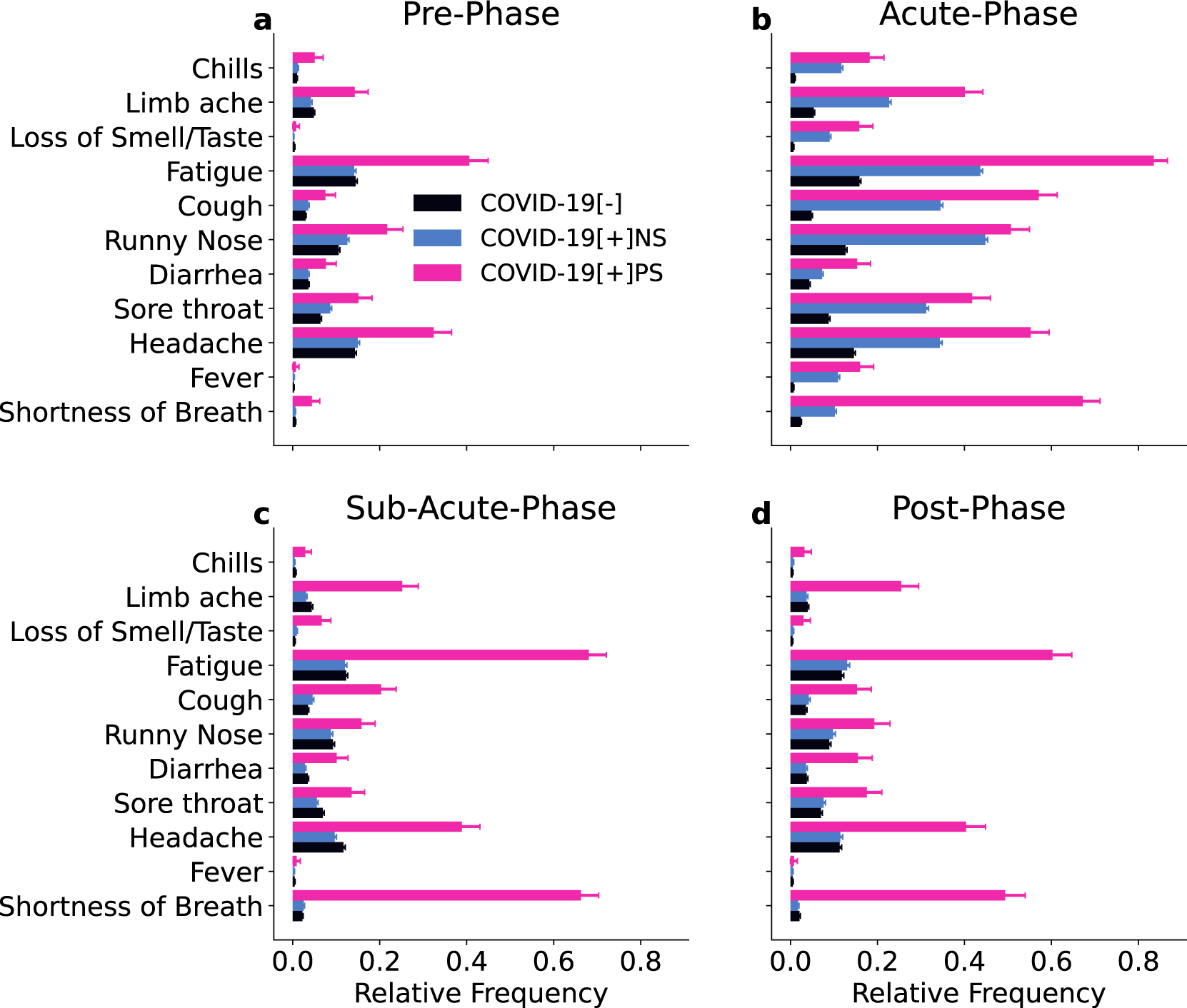
Relative frequency per symptom. For pre- (a), acute- (b), sub-acute (c) and post (d) phase we show the relative frequency per reported symptom for all three groups in the matched cohort: negative match in black (*M - COV ID -* 19[*-*]), positive match in blue (*M - COV ID -* 19[+]*NS*) and persistent symptoms match in pink (*M - COV ID -* 19[+]*PS*).

**Supplementary Figure 4.**
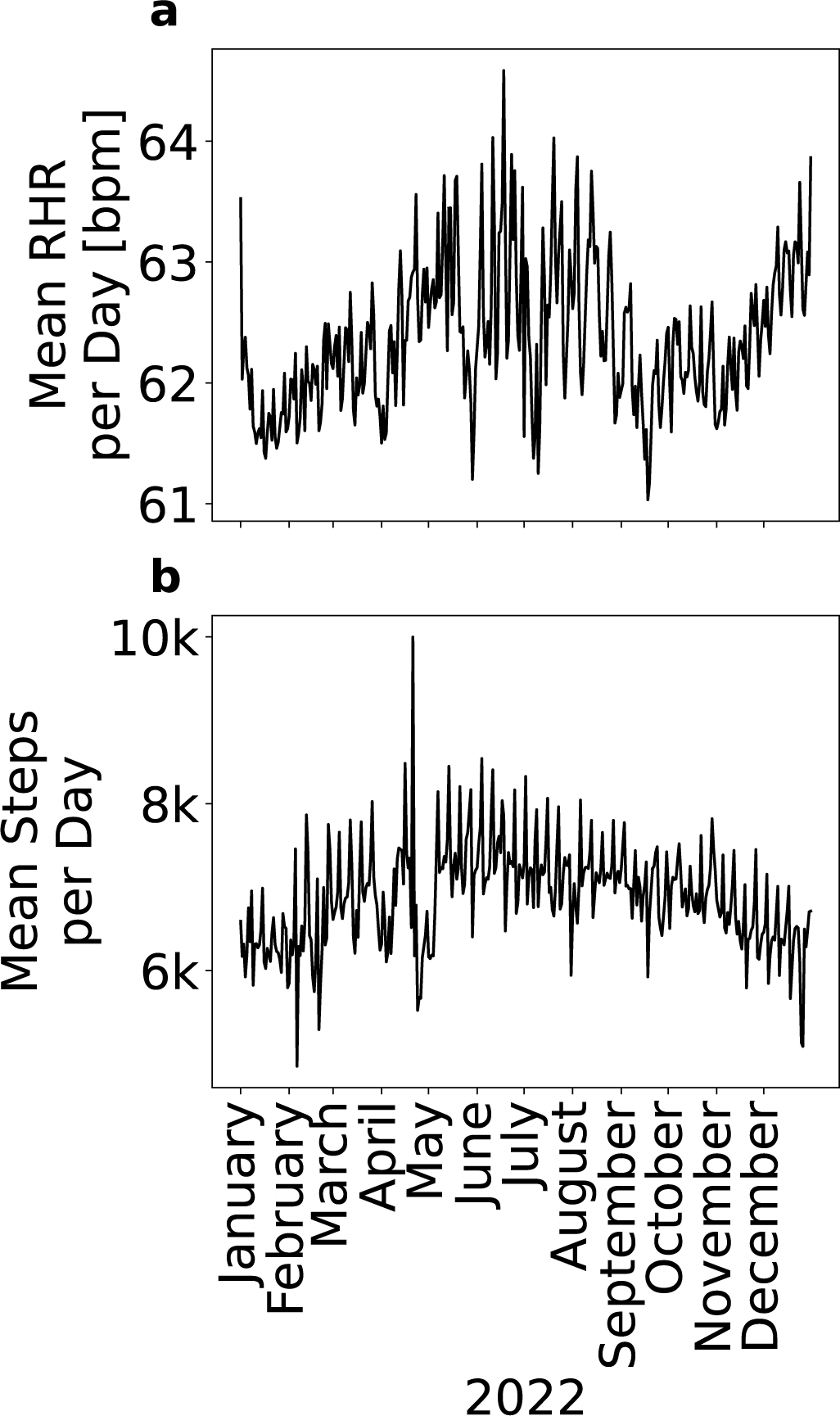
Seasonal trends in wearable data. Exemplary timeseries of mean RHR per day (a) and mean steps per day (b) over the CDA Population for one example year (2022) and one example donation-source (Apple).

**Supplementary Figure 5.**
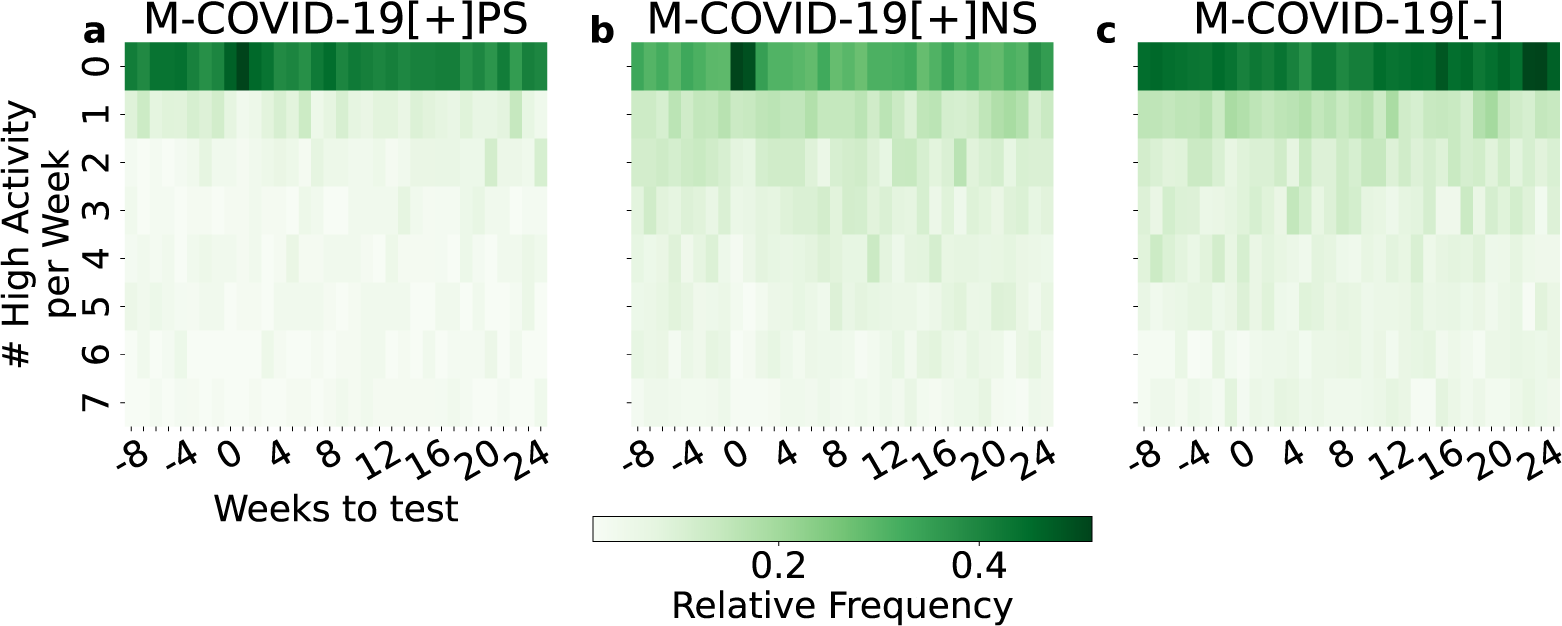
High activity profiles of the *matched cohort*. Relative frequency of the number of high activity days per week relative to the SARS-CoV-2 test date for *M - COV ID -* 19[+]*PS* (a), *M - COV ID -* 19[+]*NS* (b) and *M - COV ID -* 19[*-*] (c). A day is defined as a high activity day when the corresponding number of steps per day exceed one standard deviation over the seasonal mean taken over the whole set of users in the Corona Data Donation Project (CDA Population).

**Supplementary Figure 6.**
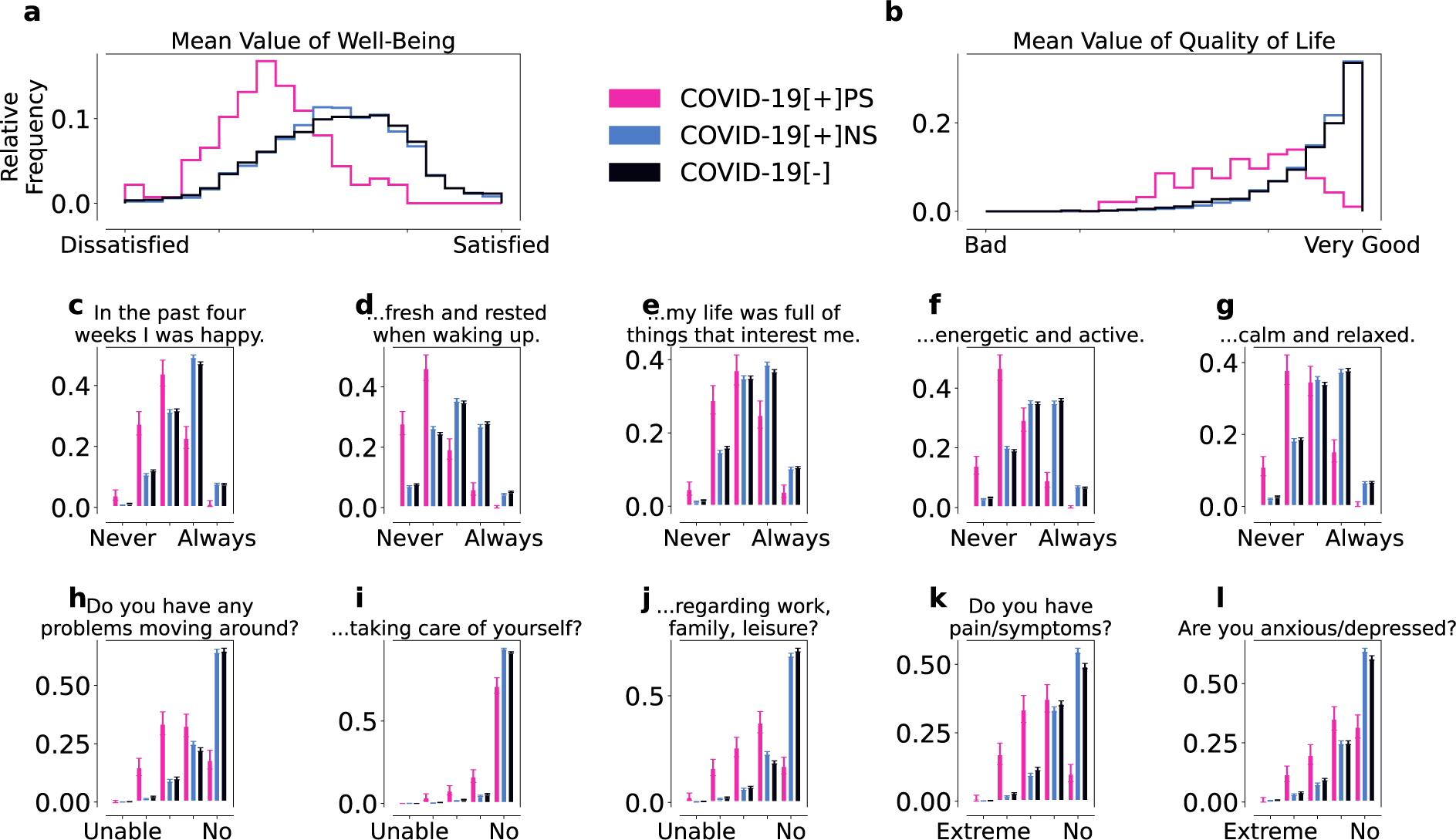
Answers to WHO-5 wellbeing and EQ-5D questionnaires before the reported SARS-CoV-2 test for the persistent symptoms (pink), positive control (blue) and negative control cohort (black). WHO-5 wellbeing (a,c-g) and modified EQ-5D/QoL (b,h-l) for the *COV ID -* 19[+]*PS* (pink), *COV ID -* 19[+]*NS* (blue), and *COV ID -* 19[*-*] (black) cohorts before the reported SARS-CoV-2 test. The individual WHO-5 and modified EQ-5D scores were both averaged to obtain the overall wellbeing (a) and QoL (b) scores, respectively. Overall, *COV ID -* 19[+]*PS* individuals reported more issues with wellbeing (c-g) and QoL (h-l) than the control cohorts. Error bars indicate standard errors.

**Supplementary Figure 7.**
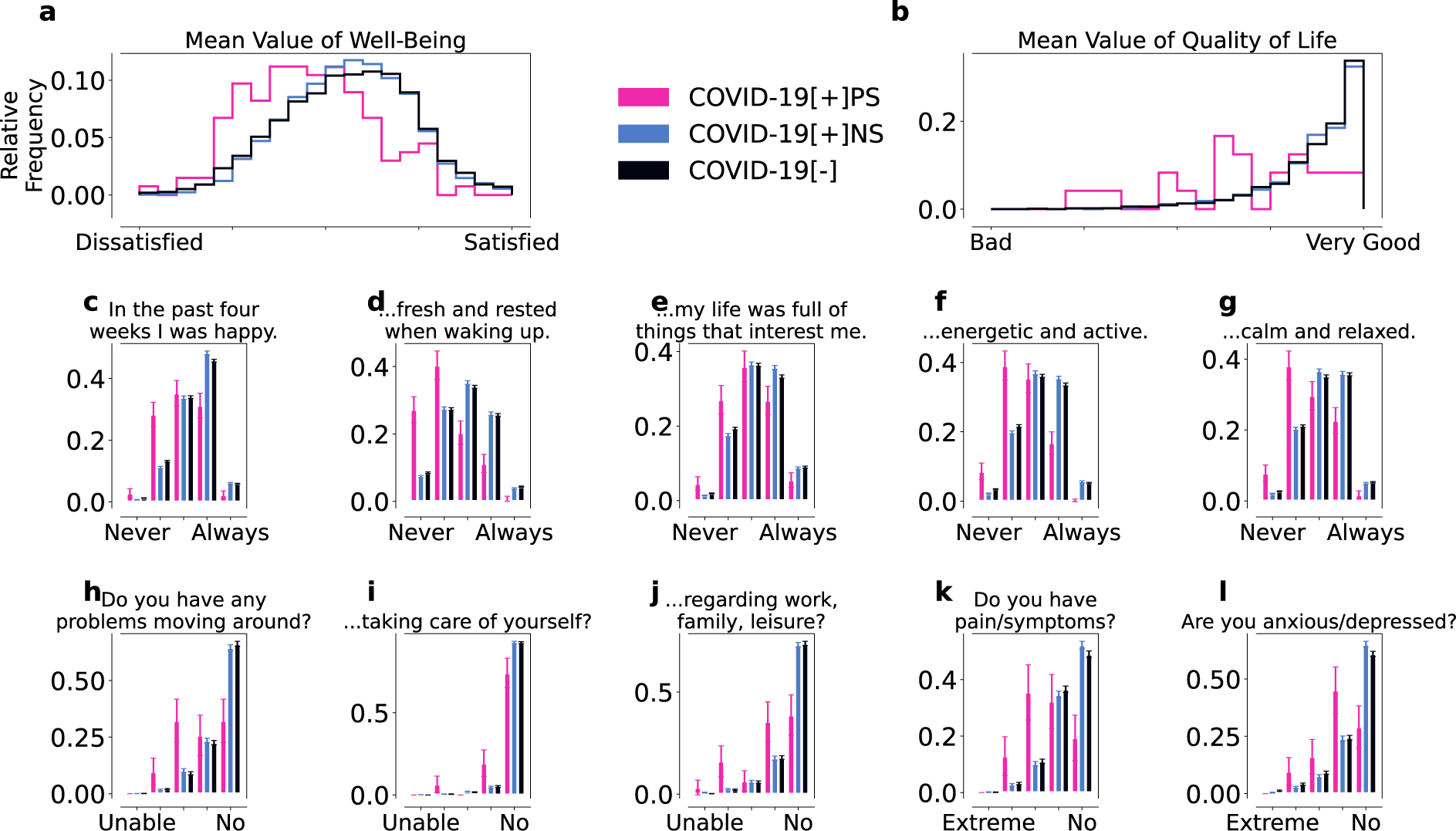
Answers to WHO-5 wellbeing and EQ-5D questionnaires after the reported SARS-CoV-2 test for the persistent symptoms (pink), positive control (blue) and negative control cohort (black). WHO-5 wellbeing (a,c-g) and modified EQ-5D/QoL (b,h-l) for the *COV ID -* 19[+]*PS* (pink), *COV ID -* 19[+]*NS* (blue), and *COV ID -* 19[*-*] (black) cohorts after the reported SARS-CoV-2 test. The individual WHO-5 and modified EQ-5D scores were both averaged to obtain the overall wellbeing (a) and QoL (b) scores, respectively. Overall, *COV ID -* 19[+]*PS* individuals reported more issues with wellbeing (c-g) and QoL (h-l) than the control cohorts. Error bars indicate standard errors.

**Supplementary Figure 8.**
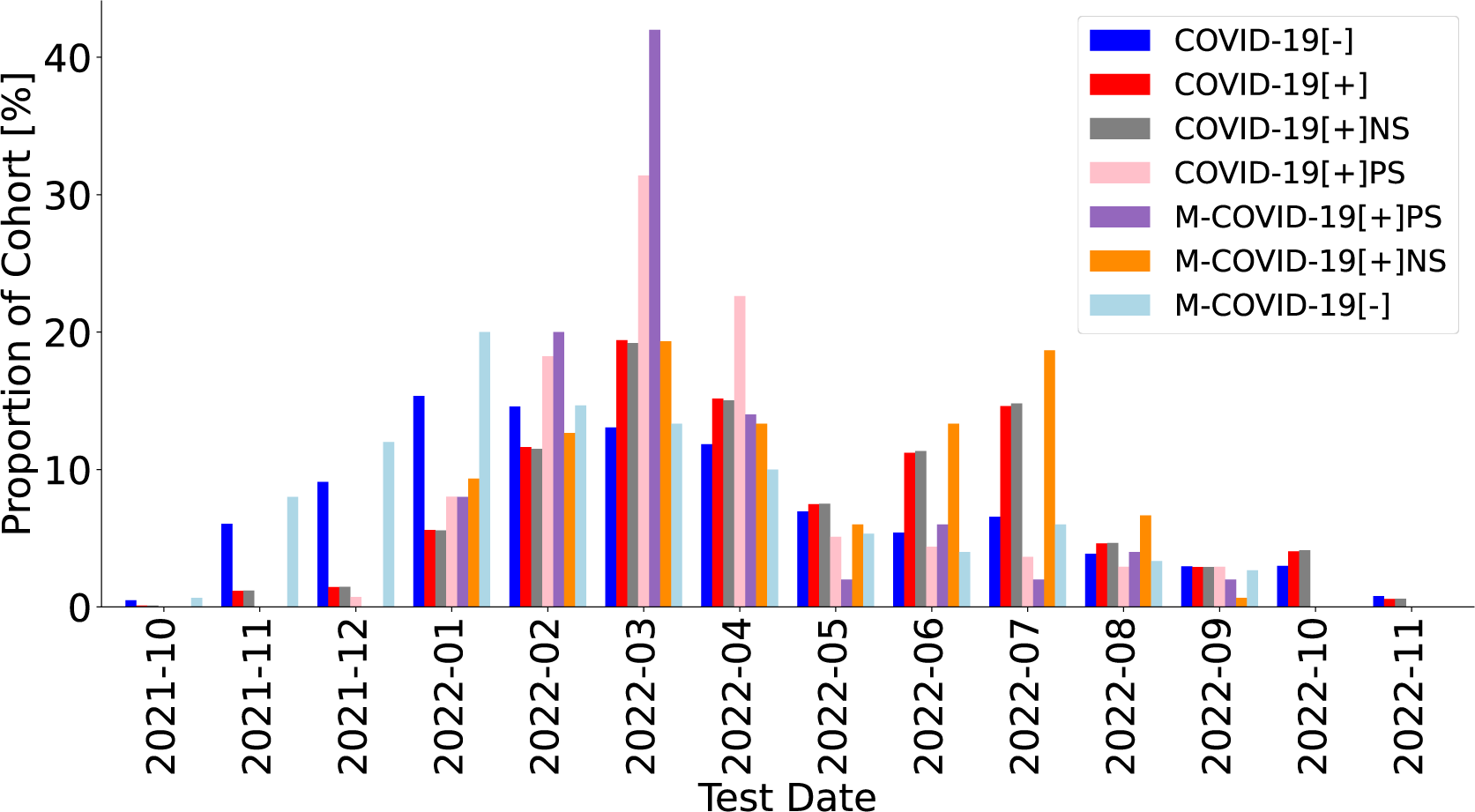
Distribution of reported SARS-CoV-2 testdates of studied cohorts. We show per month ranging from October 2021 to November 2022 the proportion of individuals who indicated their SARS-CoV-2 test in the respective month for the COVID-19[-] (negative, black), COVID-19[+] (positive, red), COVID-19[+]NS (positive control, grey), COVID-19[+]PS (persistent symptoms, pink), M-COVID-19[+]PS (persistent symptoms match, blue), M-COVID-19[+]NS (positive match, orange), and M-COVID-19[-] (negative match cohort, light black).

**Supplementary Figure 9.**
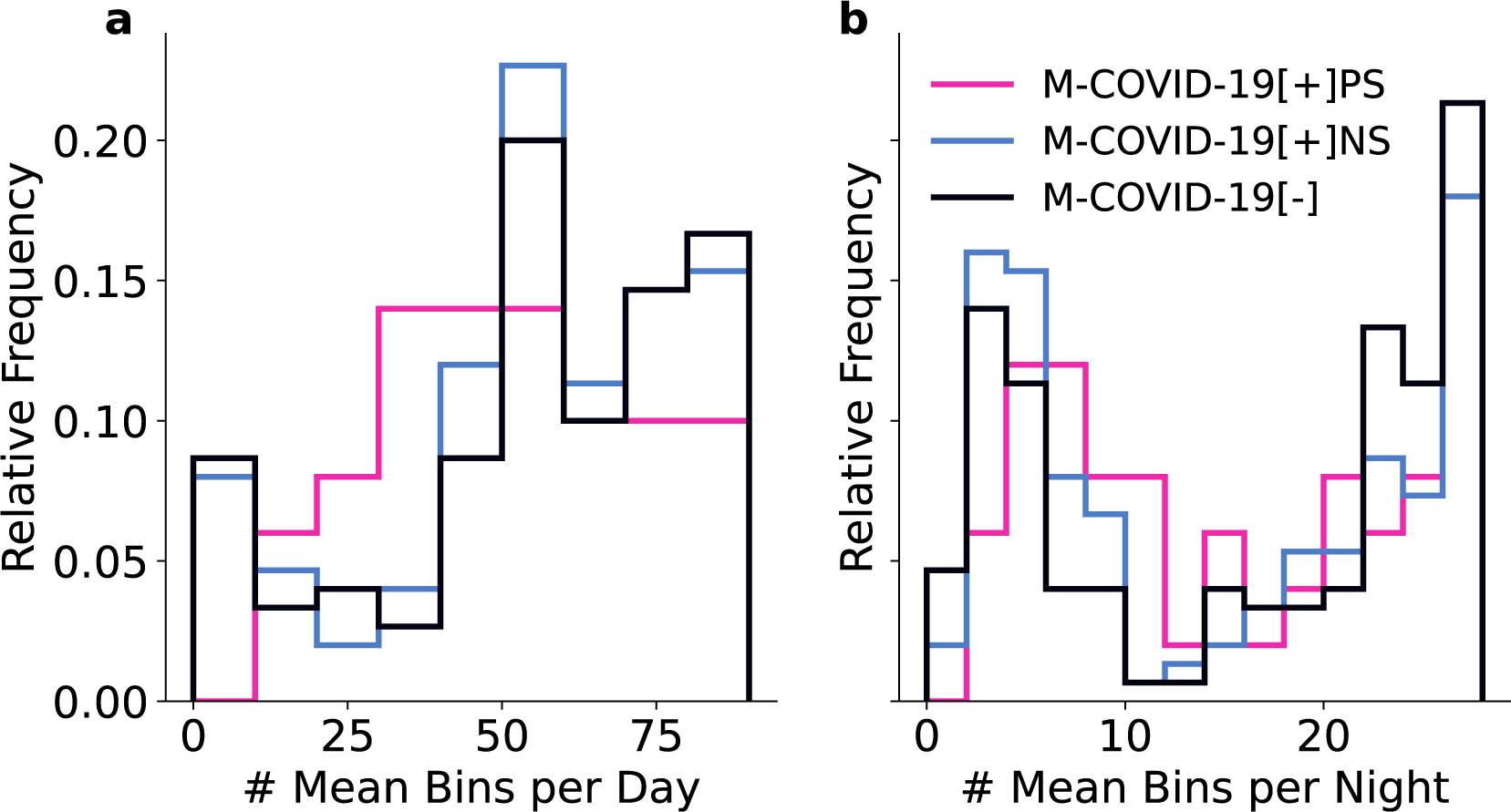
Distribution of time-worn of wearable devices. Number of 15 minute heart rate measurements as a proxy for wearing time for the three age and sex matched cohorts M-COVID-19[+]PS (persistent symptoms match, blue), M-COVID-19[+]NS (positive match, pink), and M-COVID-19[-] (negative match cohort, black) in 24 hours (a) and during night time from 11pm until 6am (b).

